# PennPRS: a centralized cloud computing platform for efficient polygenic risk score training in precision medicine

**DOI:** 10.1101/2025.02.07.25321875

**Authors:** Jin Jin, Bingxuan Li, Xiyao Wang, Xiaochen Yang, Yujue Li, Ruofan Wang, Chenglong Ye, Juan Shu, Zirui Fan, Fei Xue, Tian Ge, Marylyn D. Ritchie, Bogdan Pasaniuc, Genevieve Wojcik, Bingxin Zhao

## Abstract

Polygenic risk scores (PRS) are becoming increasingly vital for risk prediction and stratification in precision medicine. However, PRS model training presents significant challenges for broader adoption of PRS, including limited access to computational resources, difficulties in implementing advanced PRS methods, and availability and privacy concerns over individual-level genetic data. Cloud computing provides a promising solution with centralized computing and data resources. Here we introduce PennPRS (https://pennprs.org), a scalable cloud computing platform for online PRS model training in precision medicine. We developed novel pseudo-training algorithms for multiple PRS methods and ensemble approaches, enabling model training without requiring individual-level data. These methods were rigorously validated through extensive simulations and large-scale real data analyses involving over 6,000 phenotypes across various data sources. PennPRS supports online single– and multi-ancestry PRS training with seven methods, allowing users to upload their own data or query from more than 27,000 datasets in the GWAS Catalog, submit jobs, and download trained PRS models. Additionally, we applied our pseudo-training pipeline to train PRS models for over 8,000 phenotypes and made their PRS weights publicly accessible. In summary, PennPRS provides a novel cloud computing solution to improve the accessibility of PRS applications and reduce disparities in computational resources for the global PRS research community.

## Introduction

The last two decades have seen remarkable growth in genome-wide association studies (GWAS), yielding extensive data resources valuable for genetic risk prediction^1,2^. Polygenic risk scores (PRS), calculated as the sum of the number of alleles of genetic variants weighted by their effect sizes, encapsulate cumulative genome-wide risks for complex traits and diseases^3–5^. Numerous studies have highlighted the utility of PRS in precision medicine to help disease risk stratification and inform clinical intervention decisions^6–9^. To improve the accuracy and robustness of PRS, a wide range of methods, software, standards, and web resources have been developed^3,10–14^. Recent initiatives aim to further extend PRS applications to more diverse and admixed global populations^15–18^. Such efforts have been reflected by the establishment of a series of NHGRI-funded consortia, including the PRIMED^19^, which aims to develop and evaluate methods to improve the use of PRS for predicting disease risks in diverse ancestry populations, and the eMERGE Network^20,21^, which supports genomic medicine translation by returning PRS results to individuals along with healthcare recommendations in diverse clinical settings. The combination of methodological advancements, increasingly rich discovery GWAS data, and decreasing costs in biotechnology are anticipated to persistently and substantially improve both the capabilities and accessibility of PRS-based disease risk prediction and stratification.

The accessibility and scalability of PRS applications, however, are often hindered by significant challenges in the PRS model training process, particularly for users of advanced PRS algorithms (**Fig. 1a**). For example, access to high-performance computational resources required to run these algorithms and store large-scale GWAS summary data is often dependent on existing institutional infrastructure, which may not be readily available to all PRS researchers across diverse organizations and scientific fields. Additionally, managing and testing various PRS methods within local pipelines can involve a steep learning curve and make it difficult to keep up with the frequent updates to new methods. A further complication arises from the need for an independent individual-level dataset during the training process, which is typically used as tuning data for optimizing model parameters and training ensemble models. This dataset must be sufficiently large and independent from the one used to generate GWAS summary statistics. Due to privacy concerns surrounding the sharing of individual-level genetic data, obtaining such a dataset can present logistical challenges in PRS applications. Furthermore, for certain traits, even when individual-level datasets are available, their sample sizes may be insufficient to produce reliable and stable parameter tuning results.

**Fig. 1:**
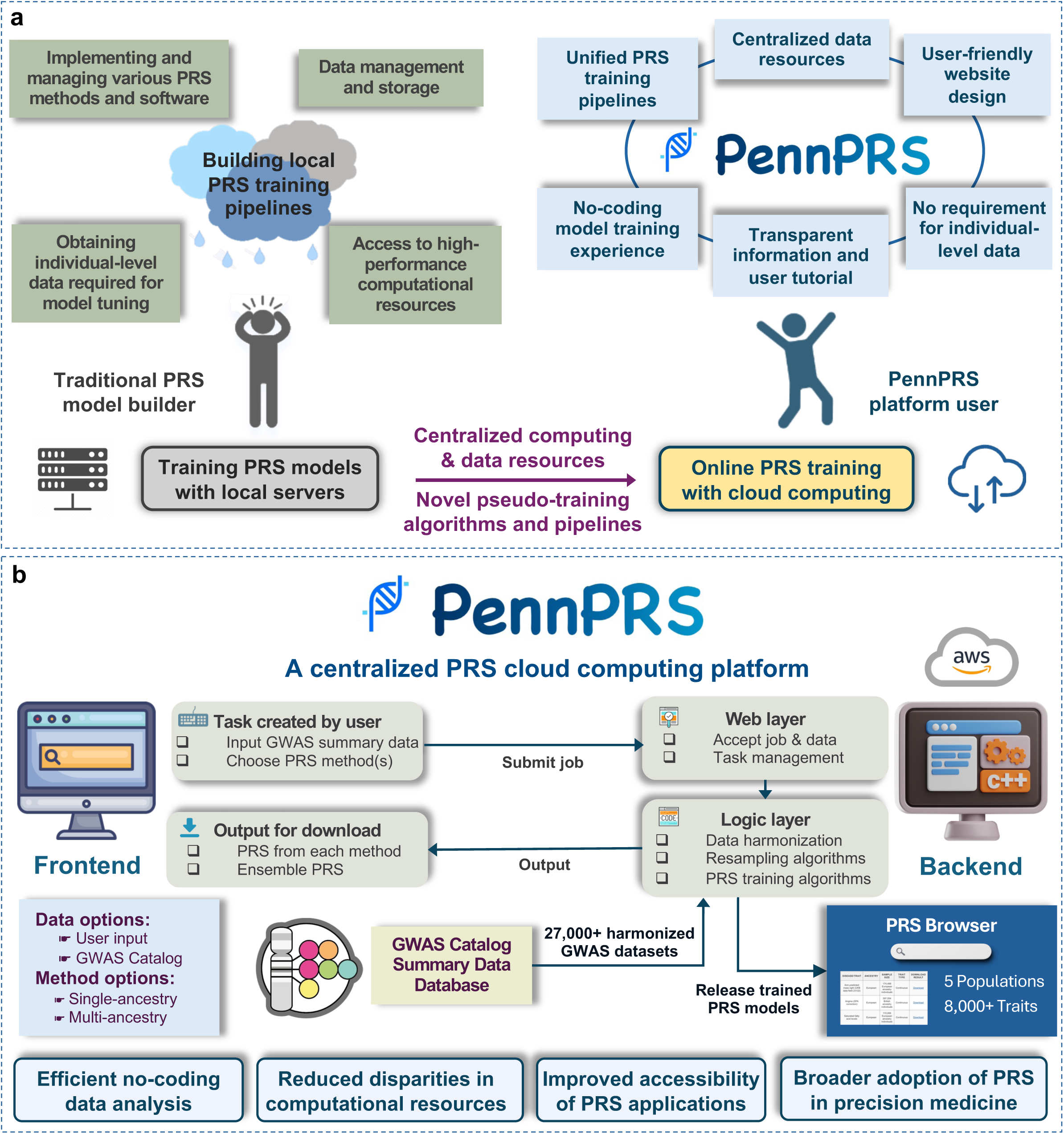
The Challenges of Traditional PRS Model Training and the Promise of PennPRS Cloud Computing Platform. **a.** Left: A figurative representation of the key challenges in performing PRS model training with local computing servers and pipelines. Right: Our proposed cloud computing approach for online PRS model training, which leverages centralized computing and data resources alongside novel pseudo-training algorithms and pipelines to overcome these challenges. **b.** An overview of the cloud computing platform of PennPRS and its major impacts on PRS applications in precision medicine.

The use of cloud computing is gaining increasing momentum in biomedical research^22–28^, especially with centralized research resources, given the energy-efficient nature of cloud computing for hosting large-scale computing and data sources with high scalability and security^29^. Several biobanks have recently developed study-centric cloud computing platforms, such as the UK Biobank Research Analysis Platform (https://ukbiobank.dnanexus.com/) and the All of Us Researcher Workbench (https://www.researchallofus.org/data-tools/workbench/), to increase their data accessibility across diverse research communities. Cloud computing provides a promising next-generation solution to address the challenges in the widespread expansion of PRS applications. By leveraging robust online resources (such as Amazon Web Services [AWS]), cloud computing can provide a well-organized platform for diverse PRS users, facilitating efficient data analysis through centralized data storage and unified pipelines. However, a key barrier to implementing PRS model training on online servers is the reliance of many PRS methods on individual-level genetic data, which raises concerns about availability and data privacy. Recent advances have introduced the pseudo-training approach for PRS model development^30–33^. This approach allows for the sampling of pseudo-training and validation summary statistics from the underlying probability distribution of GWAS summary statistics. These sampled statistics closely mimic what would be obtained if there were access to two subsets of the GWAS samples, enabling parameter tuning and the derivation of PRS models. This “self-training” approach makes it possible to generate PRS weights without the need for individual-level genetic data.

Building on these advancements, this paper aims to integrate cloud computing with pseudo-training approaches to enable online training of PRS models, providing a secure, efficient, and scalable solution for the PRS research community. We first developed pseudo-training versions for multiple single– and multi-ancestry PRS methods and rigorously showed their robust performance across thousands of phenotypes from various data sources. Based on their reliable numerical performance, we introduced PennPRS (https://pennprs.org/), a scalable cloud computing platform for online training of PRS models using summary statistics only (**Fig. 1b**). PennPRS provides a wide range of user options and supports both single– and multi-ancestry analyses across the five super populations^34^, including European (EUR), African and African American (AFR), Admixed American (AMR), East Asian (EAS), and South Asian (SAS). Users can input GWAS summary statistics, submit a job with selected methods and customized settings, and download the trained PRS models upon completion. As a centralized PRS online training platform, PennPRS provides cloud computing functionalities, extensive data resources, and offline pipelines, offering an efficient solution to PRS model development in precision medicine (**Fig. 2a**). It is designed to accommodate the training needs of various research groups with diverse requirements and computational resources.

**Fig. 2:**
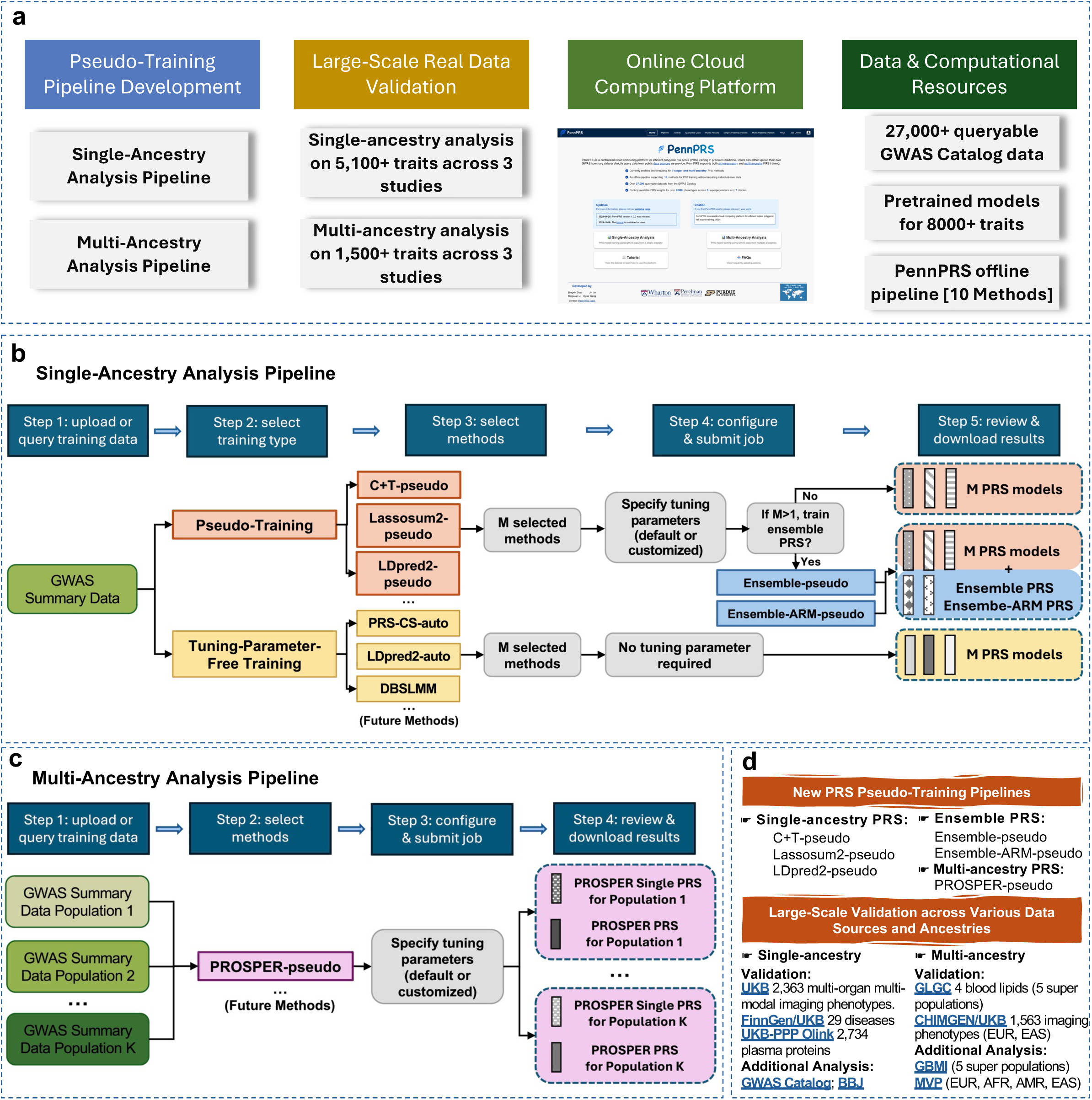
Development and Distribution of the PennPRS Cloud Computing Platform and Accompanying Data and Computational Resources. **a.** A summary of the main contributions of our study, including the distribution and large-scale validation of PRS pseudo-training pipelines, establishment of the PennPRS cloud computing platform, and distribution of queryable GWAS summary data sources, pretrained PRS models, and offline pipeline. **b.** Workflow of the single-ancestry PRS training supported by PennPRS. **c.** Workflow of the multi-ancestry PRS training supported by PennPRS. **d.** Highlighted features of PennPRS: (i) new PRS pseudo-training pipelines supporting three single-ancestry methods, two ensemble approaches combining different single-ancestry methods, and one multi-ancestry method; and (ii) large-scale application and validation of the PRS pseudo-training pipeline across nine data resources and over 6,000 phenotypes.

## Results

### Summary data-based PRS model tuning and ensemble learning

We developed single-ancestry pseudo-training pipelines with summary data-based parameter tuning for three PRS methods: Clumping and Thresholding (C+T)^4,5^, Lassosum2^35,36^, and LDpred2^37,38^, which we denote by C+T-pseudo, Lassosum2-pseudo, and LDpred2-pseudo, respectively (**Fig. 2b**). The PUMAS^31^ workflow was used to derive pseudo training and validation subsamples from the input GWAS summary statistics, enabling the selection of optimal tuning parameters. While the general framework of PUMAS pseudo-training has been established, applying it to specific PRS methods is challenging due to the complexities involved in implementing different PRS methods and non-universality of the original PUMAS software to the general GWAS summary data. Therefore, we made a series of important modifications to both the methodology and the software to ensure proper alignment between the PUMAS algorithm and each of the implemented PRS methods. For example, the original Lassosum2 and LDpred2 algorithms may generate non-convergent PRS weights under some tuning parameter settings, which can result in problematic parameter tuning. To address this potential issue, we developed a data-driven approach to improve robustness of the summary data-based parameter optimization in Lassosum2-pseudo and LDpred2-pseudo (see **Methods**).

For single-ancestry analysis, we further developed two ensemble approaches within our pseudo-training framework: Ensemble-pseudo and Ensemble-ARM-pseudo. These approaches combine PRS models trained by different methods using a linear combination strategy^39^ (Ensemble-pseudo) or an adaptive regression by mixing approach^40^ (Ensemble-ARM-pseudo) (**Fig. 2b**). The two ensemble learning methods were originally designed for use with the need for individual-level tuning datasets. Here we redeveloped them for pseudo-training within the PUMA-CUBS^31^ framework, incorporating a series of modifications to ensure robustness. Details are provided in the **Methods** section.

Notably, our pipeline supports multi-ancestry PRS training based on ancestry-stratified GWAS summary data from multiple ancestral populations, a process that is typically computationally intensive and requires more learning resources (**Fig. 2c**). We developed PROSPER-pseudo, a pseudo-training pipeline for PROSPER^41^ which is an ensemble learning-assisted multi-ancestry PRS method. PROSPER-pseudo will generate two complementary population-specific models: PROSPER-Single-pseudo and PROSPER-pseudo, where the former provides the best single PRS generated before the final ensemble step in PROSPER, and the latter provides the final PRS that combines multiple PRS across different ancestries and tuning parameter settings (see **Methods**). In summary, we have developed three single-ancestry methods, two ensemble approaches, and one multi-ancestry method as the primary methods for implementation on our cloud computing platform, all based on pseudo-training that eliminates the need for individual-level data. A summary of our pseudo-training algorithms for implementing these single– and multi-ancestry PRS methods is provided in **Supplementary Fig. 1**. Additionally, we have developed several complementary methods for the offline pipeline, which will be introduced in later sections.

### Large-scale evaluation of the PRS pseudo-training approach

We evaluated the performance of our PRS pseudo-training methods through extensive simulations and real data analyses. Simulation results of the single-ancestry methods under various settings of genetic architecture of the phenotype (such as heritability and causal variant proportion) and GWAS sample size^42^ demonstrate that our PRS pseudo-training methods perform comparably to those original methods that tuned model parameters with a sufficiently large hold-out individual-level dataset (e.g., *N_tuning_* = 1000, **Fig. 3a, Supplementary Fig. 2**, and **Supplementary Table 1**). As training GWAS sample size increases, our pseudo-training methods tend to better approximate the PRS under the optimal tuning parameter setting. Pseudo-training versions of the ensemble PRS tend to have slightly lower prediction R-squared (*R*^2^) than individual-level data-based ensemble. It is important to note that the above comparisons assume access to sufficiently large individual-level tuning data. However, when the number of individual-level tuning samples is insufficient (e.g., *N_tuning_* < 1,000), pseudo PRS training notably outperforms traditional PRS training methods that rely on individual-level tuning data (**Figs. 3b** and **3c**). For example, compared to the traditional PRS training pipelines using an individual-level tuning dataset of size *N_tuning_* = 400 or 100, our PRS pseudo-training pipeline for the same PRS methods achieved a 6.5% or 44.7% higher *R*^2^, respectively. These findings highlight the utility of the PRS pseudo-training methods we developed, particularly in scenarios where individual-level data is limited or unavailable for parameter tuning.

**Fig. 3:**
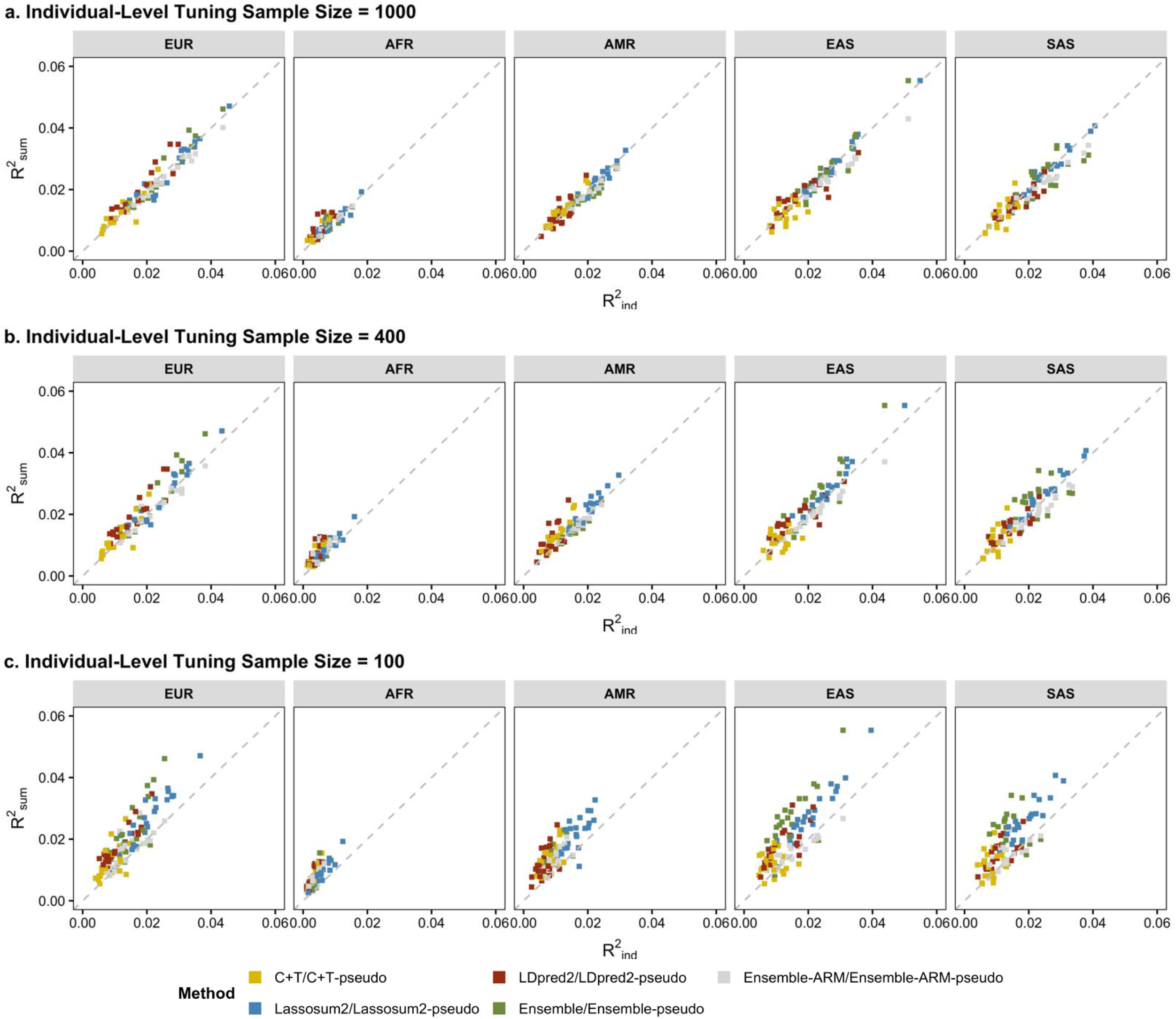
Comparison of Single-ancestry PRS Pseudo-training and Traditional PRS methods with Individual-level Tuning Data of Various Sample Sizes under various settings of causal SNP proportion and heritability. We compared the prediction *R*^2^ of the PRS models trained by C+T-pseudo, Lassosum2-pseudo, LDpred2-pseudo, Ensemble-pseudo, and Ensemble-ARM-pseudo (*R_2sum_*) with those of PRS models trained based on individual-level tuning dataset (*R^2^_ind_*) that has a sample size **a**. *N_tuning_*=1,000, **b.** *N_tuning_*=400, or **c.** *N_tuning_*=100. Results were summarized across 10 training GWAS summary datasets of *N_GWAS_*=15,000 and averaged across 100 random splits, with each split having *N_tuning_* tuning samples for individual data-based parameter tuning and *N_val_*=2,500 validation samples for calculating prediction *R*^2^ for all models. Detailed results are reported in Supplementary Table 1.

We examined the performance of our pseudo-training pipelines for single-ancestry PRS model training across different phenotypes and data sources (**Fig. 2d**). First, we used 2,106 multi-organ multi-modal imaging phenotypes with GWAS summary data available from the UK Biobank (UKB)^43^ study, including 1,432 brain structural magnetic resonance imaging (sMRI)^44^, 674 diffusion MRI (dMRI), 82 resting-state functional MRI (rfMRI), 41 abdominal MRI^45^, 82 cardiovascular MRI^46^, and 46 eye optical coherence tomography images (OCT)^47^ (average *N* = 32,859). These imaging phenotypes are well-established clinical biomarkers with widespread practical applications in precision medicine^48^. We found that all three single-ancestry pseudo-training methods, as well as the two pseudo-training ensemble approaches, demonstrated strong performance across these diverse imaging modalities (**Figs. 4a** and **5a, Supplementary Tables 2-5**; mean *R*^2^ = 0.0562 vs. 0.0567, *R*^2^ correlation = 0.955). Consistent with our simulation studies, we observed that pseudo-training methods outperform individual-level data-based tuning as the individual-level tuning sample size decreases (**Fig. 4b** and **Supplementary Fig. 3**). For example, with a tuning sample size *N_tuning_* = 1,000, 300, and 100, our pseudo-training methods produced PRS with *R*^2^ values that were 0.4%, 6.9%, and 18.5% higher, respectively, compared to methods using individual-level tuning data for eye OCT phenotypes (**Fig. 4b**).

**Fig. 4:**
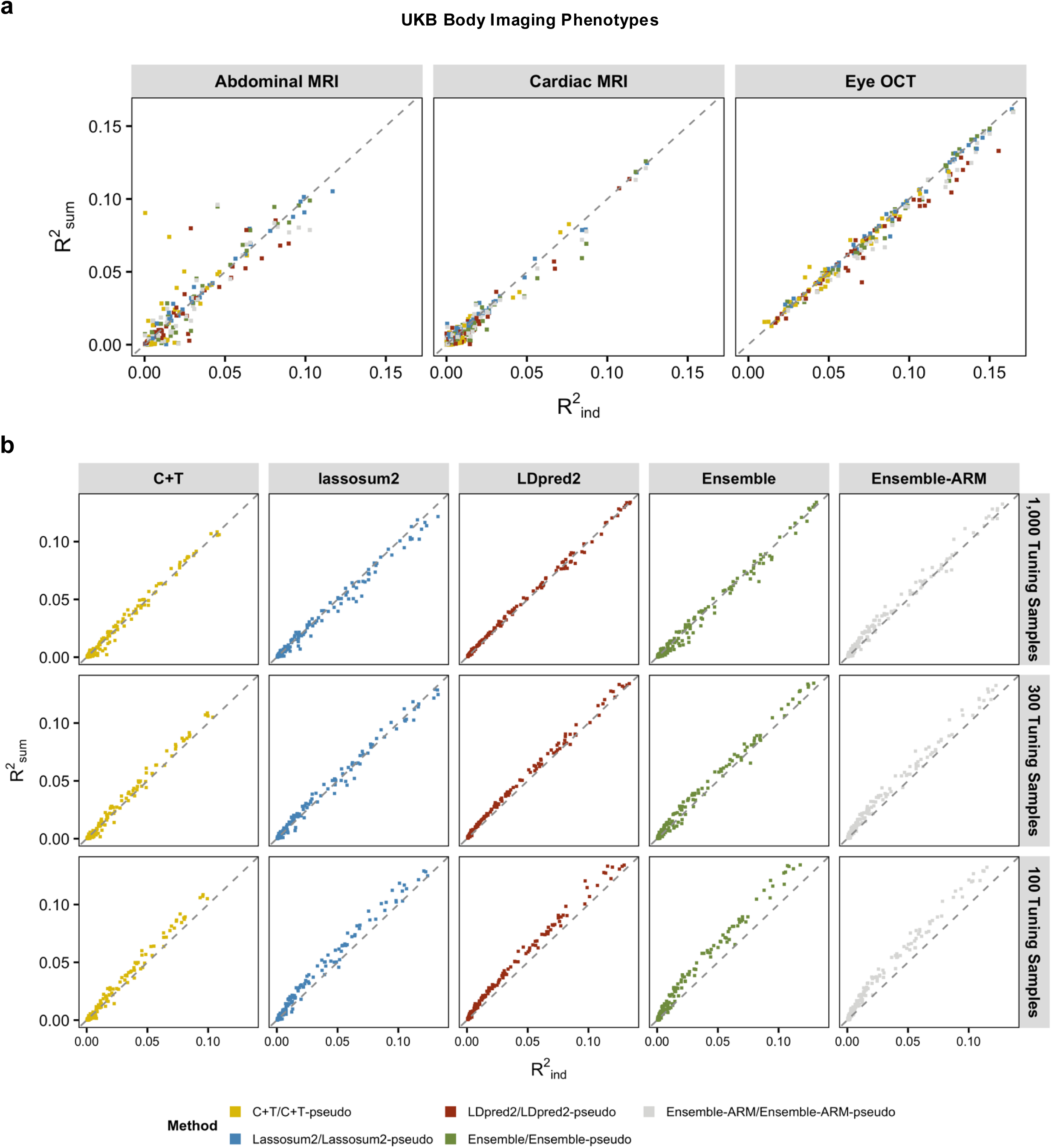
Evaluation of Single-ancestry PRS Pseudo-training on Body Imaging Phenotypes Using GWAS Summary Data and Validation Data from the UK Biobank (UKB) study. **a.** We compared our PRS pseudo training approaches, C+T-pseudo, Lassosum2-pseudo, LDpred2-pseudo, Ensemble-pseudo, and Ensemble-ARM-pseudo (*R^2^_sum_*) with the original methods that use individual-level tuning dataset (*R^2^_ind_*) on 41 abdominal MRI (average *N_GWAS_*=29,849), 82 cardiac MRI (average *N_GWAS_*=30,506), and 46 eye OCT (average *N_GWAS_*=50,465) phenotypes and evaluated their performance on hold-out independent UKB samples of EUR origin (*N_val_*=5,760). **b.** We assessed the relative performance of the pseudo-training methods to their original versions utilizing individual-level tuning datasets of different sizes *N_tuning_*= 1,000, 300, or 100, on the abdominal MRI, cardiac MRI, and eye OCT phenotypes. Results were averaged across 100 random splits, with each split having *N_tuning_* tuning samples for individual data-based parameter tuning and the remaining samples for calculating prediction *R*^2^ for all models. Detailed data information and results are summarized in Supplementary Tables 2-3.

**Fig. 5:**
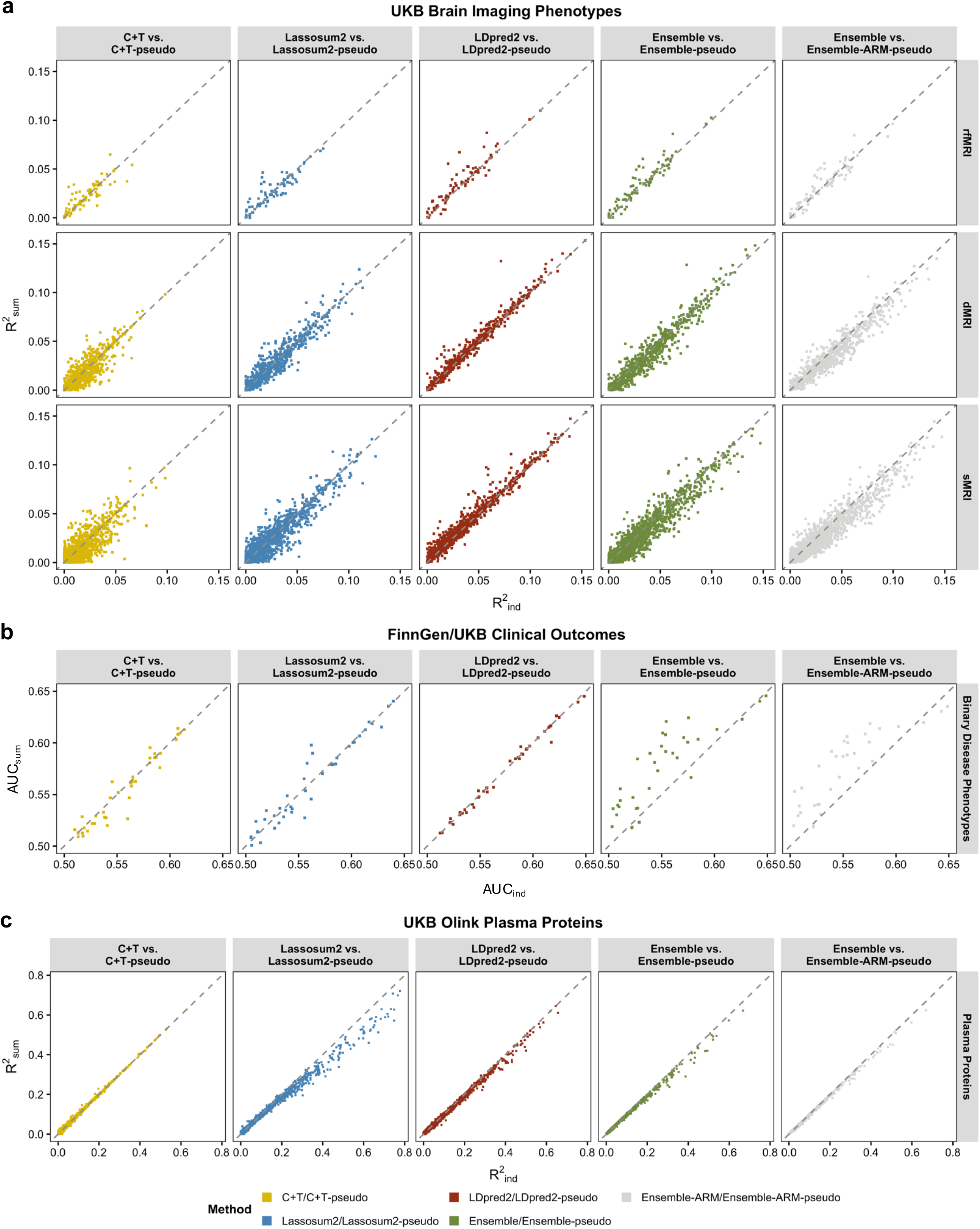
Additional Evaluation of Single-ancestry PRS Pseudo-training across Various Phenotypes and Data Sources. We compared our PRS pseudo-training approaches, C+T-pseudo, Lassosum2-pseudo, LDpred2-pseudo, Ensemble-pseudo, and Ensemble-ARM-pseudo (*R^2^_sum_*) with the original methods that use individual-level tuning dataset (*R^2^_ind_*) on **a**. 2,363 brain multi-modal imaging phenotypes based on GWAS summary statistics of EUR ancestry from the UK Biobank (UKB) study (*N_GWAS_*=32,620) and evaluated their performance on hold-out independent UKB samples of EUR ancestry (*N_val_*=5,020); **b.** 29 binary disease phenotypes based on GWAS summary statistics of EUR ancestry from the FinnGen study (a total of 333,355 cases and controls on average) and evaluated their performance on UKB samples of EUR ancestry (23,048 cases on average); and **c.** 2,734 Olink plasma proteins based on GWAS summary statistics of EUR ancestry from the UKB-PPP project (*N_GWAS_*=40,852) and evaluated their performance on hold-out independent UKB samples of EUR ancestry (*N_val_*=2,731). We used half of the UKB validation samples for individual-level parameter tuning and the remaining half to report AUC (for binary disease phenotypes) and prediction *R*^2^ (for continuous phenotypes) for both our pseudo-training approach and the individual-level tuning data-based training approach. Here rfMRI stands for resting-state functional MRI, dMRI stands for diffusion MRI, and sMRI stands for structural MRI. Detailed data information and results are summarized in Supplementary Tables 4-9.

Furthermore, we trained PRS for 29 binary disease phenotypes based on GWAS summary statistics from the FinnGen^49^ study and evaluated their performance on matched clinical outcomes using UKB testing individuals^50^ (average *N_case_* = 23,048, **Supplementary Table 6**). Again, all three single-ancestry pseudo-training methods demonstrated performance comparable to the traditional methods using individual-level tuning data (area under the ROC curve [AUC] correlations = 0.880). Notably, the pseudo-training ensembles outperformed the individual-level data ensembles, even though the individual-level tuning datasets are large (**Fig. 5b** and **Supplementary Table 7**; mean AUC = 0.564 vs. 0.555, one-sided *P* = 1.51×10^-7^). In addition, we evaluated the PRS performance on 2,734 Olink plasma proteins with GWAS data from the UKB-PPP project^51^ (average *N* = 40,852, **Fig. 5c** and **Supplementary Tables 8-9**). Plasma proteins, which are crucial for disease diagnosis and treatment^52,53^, exhibit a unique special architecture, generally showing higher heritability and with *cis*-loci accounting for a large proportion of genetic variation^51^. For such genetic architecture, our analysis revealed that C+T-pseudo and LDpred2-pseudo showed highly consistent performance with training based on individual-level tuning data (mean *R*^2^ = 0.0562 vs. 0.0567, *R*^2^ correlation = 0.998), whereas Lassosum2-pseudo consistently delivered sub-optimal performance for proteins with high prediction *R*^2^ (e.g., > 0.40) (mean *R*^2^ = 0.475 vs. 0.557). These findings suggest that C+T-pseudo and LDpred2-pseudo may be more suitable for deriving genetic scores^53^ for these proteins and other molecular traits with similar genetic architecture.

For multi-ancestry PRS training, our simulation studies suggested that PROSPER-Single-pseudo, the pseudo-trained best single PROSPER PRS without implementing the final ensemble step, approximates its individual-level data-based version (PROSPER-Single) well, while the PROSPER-pseudo PRS (with the final ensemble step) tends to perform slightly worse than PROSPER PRS, its individual-level data-based version, if large hold-out individual-level dataset exists (**Fig. 6a** and **Supplementary Table 10**). We further evaluated their performance in multi-ancestry real data applications using ancestry-stratified GWAS summary statistics (**Supplementary Tables 11-14**). We first analyzed four blood lipids, including high-density lipoprotein (HDL), low-density lipoprotein (LDL), log-transformed triglycerides (logTG), and total cholesterol (TC). We used GWAS summary data for EUR, AFR, AMR, EAS, and SAS from the Global Lipids Genetics Consortium^54^ (GLGC) (*N* = 33,658-930,671). The performance was evaluated on UKB validation individuals of EUR, AFR, AMR, EAS, and SAS ancestries^54,55^. Our results showed that pseudo-training had consistent performance across all ancestries (**Fig. 6b**, mean *R*^2^: 0.084 [PROSPER-Single-pseudo] and 0.088 [PROSPER-pseudo] vs. 0.089 [PROSPER], *R*^2^ correlation = 0.93 and 0.95, respectively). We also evaluated the performance of multi-ancestry pseudo-training using GWAS summary statistics for 1,413 brain dMRI and sMRI phenotypes from the Chinese Imaging Genetics (CHIMGEN) study^56^, jointly with matched imaging phenotypes in the UKB study. Specifically, the inputs were the CHIMGEN summary statistics (average *N* = 7,058) and UKB European summary statistics (average *N* = 32,620), with performance evaluated in independent testing data from hold-out UKB samples (average *N* = 2,510 for EUR ancestry and *N* = 222 for EAS ancestry). We found that pseudo-training outperformed individual-level data training for both EUR (*R*^2^ = 0.027 vs. 0.023), which has sufficient tuning samples, and EAS (*R*^2^ = 0.010 vs. 0.008), which has limited tuning samples, although the results for EAS had larger uncertainty due to the much smaller testing sample sizes (**Fig. 6b** and **Supplementary Fig. 4**). As expected, analyses of GLGC and CHIMGEN data also showed improved prediction accuracy when GWAS training data from both ancestries were integrated (**Fig. 6c**) and the consistent pattern of relative performance of PROSPER-Single-pseudo and PROSPER-pseudo across different data resources (**Supplementary Fig. 5**).

**Fig. 6:**
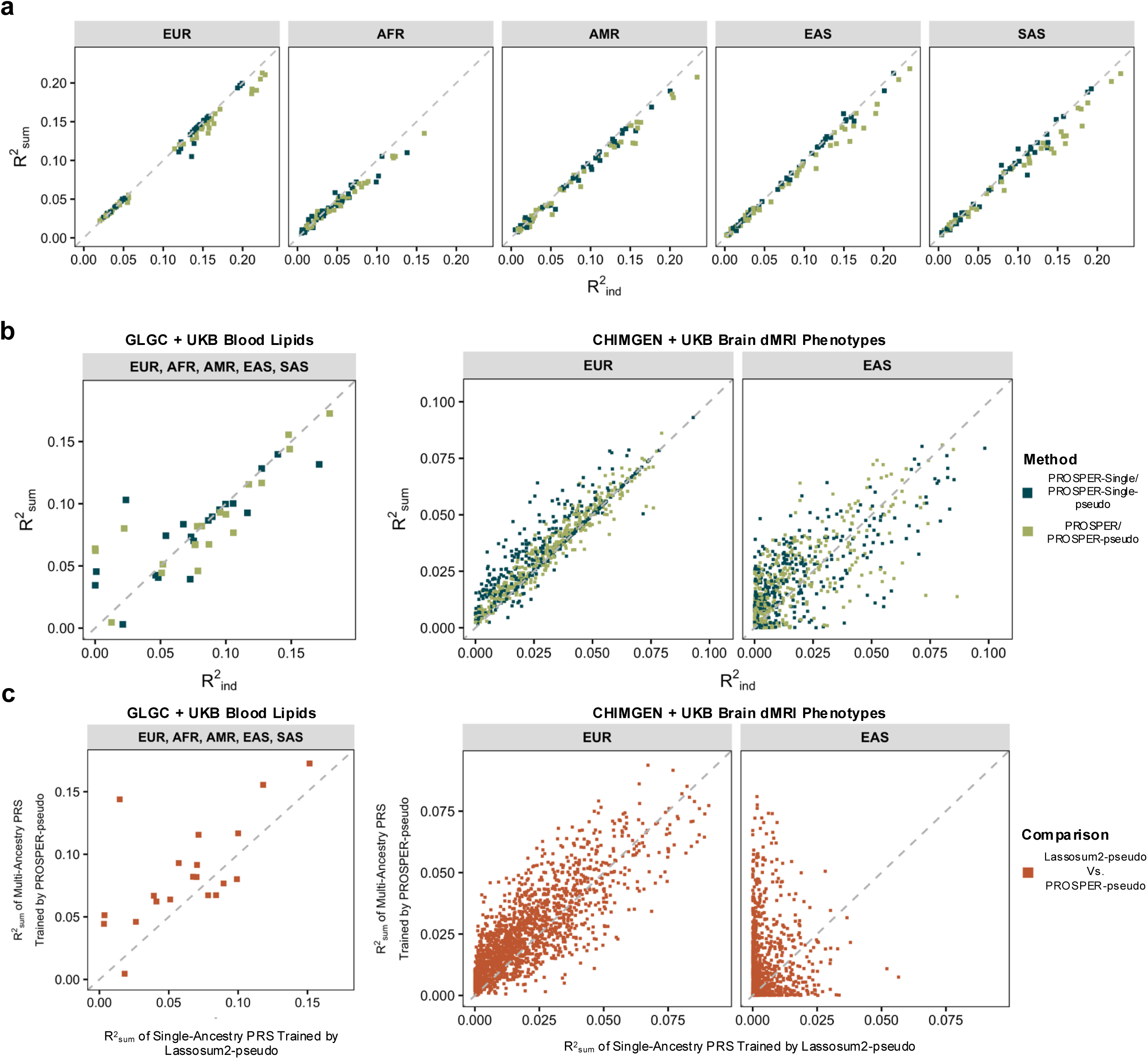
Evaluation of Multi-ancestry PRS Pseudo-training by Simulation Studies and Applications on Various Phenotypes and Data Sources. **a.** Results show the comparison of the PRS trained by pseudo-training methods (*R^2^_sum_*, PROSPER-Single-pseudo and PROSPER-pseudo) with PROSPER-Single PRS and PROSPER PRS trained with individual-level tuning datasets (*R^2^_ind_*) on **a**. simulated datasets under different settings of heritability, negative selection patterns, and causal genetic variant proportions assuming a 100,000 GWAS sample size for EUR and varying GWAS sample sizes for each non-EUR population (15,000, 45,000, or 80,000) with 2,500 tuning samples for individual data-based parameter tuning and 2,500 validation samples for calculating prediction *R*^2^ for the various models; **b**. Four blood lipid phenotypes based on GWAS summary statistics of EUR, AFR, AMR, EAS, and SAS ancestries from the GLGC study (*N_GWAS_*=33,658-930,671) and evaluated their performance on independent UK Biobank (UKB) samples (*N_val_*=1,752-19,030); and 382 brain diffusion MRI (dMRI) phenotypes based on GWAS summary data of EUR ancestry from the UKB study (*N*_GWAS_=28,626-32,744) and GWAS summary data of EAS ancestry from the CHIMGEN study (*N_GWAS_*=7,058) and evaluated their performance on hold-out independent UKB samples (*N_val_*=4,955 for EUR and *N_val_*=413-444 for EAS, half for parameter tuning for the original PROSPER method and the remaining half for calculating *R*^2^ for all models). **c.** Comparison of the performance of multi-ancestry method, PROSPER-pseudo, and its single-ancestry analogue, Lassosum2-pseudo, on the 382 dMRI phenotypes. Detailed data information and results are summarized in Supplementary Tables 11-14.

Overall, our large-scale numerical results demonstrate that, without access to an independent individual-level tuning dataset, the developed summary data-only pseudo-training methods can produce PRS weights comparable to those generated using a large individual-level tuning dataset. Furthermore, pseudo-training may even outperform traditional methods, particularly when the tuning dataset is limited in size. These findings lay the methodological groundwork for the development of PennPRS as a centralized cloud computing solution for online PRS model training.

### PennPRS: a cloud computing platform for the global PRS community

We established a centralized cloud computing platform hosted on AWS to implement the developed pseudo-training methods, enabling users to freely train PRS weights online using GWAS summary statistics (**Fig. 1b**). Upon completing registration, users can upload GWAS summary statistics or query over 27,000 harmonized summary statistics from the GWAS Catalog^57^, select PRS methods and model parameters, and submit jobs. These jobs are managed by the queuing system and processed on our server, and users can download the generated PRS weights and log files once the job is completed. In addition to the set of newly developed pseudo-training PRS methods (C+T-pseudo, Lassosum2-pseudo, LDpred2-pseudo, and PROSPER-pseudo) and pseudo-training ensemble methods (Ensemble-pseudo and Ensemble-ARM-pseudo), PennPRS supports three existing tuning-parameter-free methods (PRS-CS-auto^58^, LDpred2-auto^37^, and DBSLMM^59^). Our platform presents a robust frontend-to-backend web infrastructure with detailed tutorials and a comprehensive data harmonization pipeline to ensure regularized PRS training and an efficient user experience (see **Methods**).

Similar to many biomedical cloud computing platforms in other fields^22–27^, the PennPRS team will cover data analysis expenses for all users. This approach aligns with our mission to make PRS accessible to more researchers and, ultimately, to study participants in precision medicine, while reducing disparities in computational resources within the global PRS research community and the broader fields of genetic and medical research. To optimize the efficiency of our computational infrastructure, we conducted various tests to determine the optimal configurations for our platform, such as RAM and CPU deployment for different PRS methods implemented. We also conducted extensive tests to validate the platform’s stability and computational performance. For example, using the three single-ancestry pseudo-training methods and their two ensemble approaches as a case study, we analyzed the runtime for each step in the algorithm. We found that using 2 CPUs (with 30 GB RAM) allowed a typical job to complete in approximately two and a half hours, while increasing to 4 CPUs reduced the runtime to around two hours (**Supplementary Table 15**). Based on these empirical observations, we have optimized our configuration to make efficient use of AWS resources, currently supporting up to eight concurrent user jobs. The AWS cloud computing service, additionally supported by our local computing IT teams, provides a flexible management system for CPU and RAM, enabling us to easily maintain the server and adjust resource allocation for scaling up or down as needed.

### Public sources: GWAS Catalog data querying and working examples

The GWAS Catalog^57^ has become an invaluable database of public GWAS summary statistics, with a fast-growing collection of data curated and harmonized for post-GWAS applications. We have developed a feature that links PennPRS directly to the GWAS Catalog database to enable efficient PRS model training. This allows users to query data from the GWAS Catalog for PRS pseudo training directly without the need to download, preprocess, and upload the data to PennPRS. To enable this functionality with high quality, we focused on harmonized datasets from the GWAS Catalog and ensured that the provided data meet the basic requirements for implementing the various PRS methods supported, such as having the necessary GWAS summary-level data information and excluding data from exome studies. Currently, we provide access to over 27,000 datasets for users to query directly through PennPRS.

We provide two examples of querying GWAS Catalog datasets for efficient PRS pseudo-training on PennPRS. The first example demonstrates the use of the PennPRS single-ancestry data analysis pipeline to train a PRS model for height in Hispanics. In this example, we first navigated to the PennPRS GWAS Queryable Database (https://pennprs.org/data) and searched for “height”. We identified the dataset from the study “GCST90095033” as a suitable input for PRS training, which provided GWAS summary statistics from 59,771 Hispanic or Latin American individuals^60^. We then created a single-ancestry data analysis job on PennPRS, entering the relevant dataset information (e.g., study accession ID) to enable direct querying from the GWAS Catalog. Next, we selected three pseudo-training methods (C+T-pseudo, Lassosum2-pseudo, and LDpred2-pseudo) along with the ensemble option, which would utilize two ensemble methods, Ensemble-pseudo and Ensemble-ARM-pseudo, to train two ensemble PRS models combining PRS trained by the three selected methods and submitted the job. PennPRS completed the job in approximately two and a half hours, returning five PRS models along with a detailed log file. Similarly, the second example demonstrates the use of the PennPRS multi-ancestry data analysis pipeline to train PRS models for height across four ancestries (EUR, AFR, AMR, and EAS). We queried four corresponding ancestry-specific GWAS datasets from the GWAS Catalog (“GCST90029008”, “GCST90013468”, “GCST90095033”, and “GCST90018739”) and used the multi-ancestry method, PROSPER-pseudo, for online training. PennPRS completed this job in approximately ten and a half hours, generating eight PRS models for the four ancestries (PROSPER-Single-pseudo PRS and PROSPER-pseudo PRS for each ancestry). Detailed steps and illustrations for these examples are available in the tutorial (https://pennprs.gitbook.io/pennprs), serving as quick-start guides for new users.

### PennPRS offline pipeline and pretrained PRS models

In addition to establishing the online PRS training server, we have developed a comprehensive pipeline for offline implementation of the supported PRS pseudo-training and tuning-parameter-free methods. The cloud computing server is designed as a convenient and eco-friendly^29^ online tool for PRS users, particularly those who face challenges in setting up local computational clusters, while the offline pipeline is powerful for large-scale analyses if researchers have access to high-performance computing clusters. In our offline pipeline, we have additionally developed novel pseudo-training versions of three single– and multi-ancestry PRS methods, including PRS-CS-grid^58^-pseudo, PRS-CSx^61^-pseudo, and MUSSEL^55^-pseudo. Due to the nature of these methods, they have much higher memory and computational demands than other methods and are therefore included exclusively in our offline pipeline. By offering both online and offline options, we aim to accommodate the diverse needs of research groups and help reduce disparities in computational resources for the PRS application community.

To demonstrate the power and efficiency of our offline pipeline, we applied it to a wide range of phenotypes, including those mentioned in our model evaluations (for which we had access to individual-level testing data for performance assessment), as well as many more GWAS summary datasets from the GWAS Catalog^57^, Biobank Japan (BBJ)^62^, the Million Veteran Program (MVP) study^63^, and the Global Biobank Meta-analysis Initiative (GBMI) consortium^64^. Specifically, we have conducted single-ancestry analysis with all three single-ancestry pseudo-training methods (C+T-pseudo, Lassosum2-pseudo, and LDpred2-pseudo) and their ensembles (Ensemble-pseudo and Ensemble-ARM-pseudo) using default tuning parameter settings on over 8,000 harmonized GWAS Catalog datasets and 169 phenotypes from BBJ. We have also conducted multi-ancestry analysis with PROSPER-pseudo on 181 ancestry-stratified GWAS summary datasets from the MVP and nine ancestry-stratified GWAS summary datasets from the GBMI. We have made these pretrained PRS models publicly available in the PennPRS public resource hub (https://pennprs.org/result), allowing users to freely download and utilize them in their research. As the GWAS Catalog and other databases continue to expand, harmonizing and making more curated GWAS summary statistics publicly available, we will leverage the established PennPRS pipeline to analyze these summary datasets and share the trained PRS models with the PRS research community. These resources will accelerate the application of PRS across various fields.

## Discussion

PRS training methods and their cluster-based implementations have been traditionally handled by local servers, typically established by individual research groups. However, the fast-paced evolution of PRS methodologies, along with the growing volume of GWAS resources, presents logistical, computational, and environmental challenges for hosting these PRS pipelines locally. This is particularly true for smaller research groups that may lack sufficient computational resources or are new to PRS. In this paper, we developed a series of pseudo-training algorithms, data resources, and cloud computing functionalities to enable online single– and multi-ancestry PRS pseudo-training using summary data only, eliminating the need for local setups. Our platform aims to lower the barriers to PRS applications across various phenotypes and ancestry populations, while also reducing disparities in computational resources within the global PRS research community.

The development of cloud computing platforms and centralized resources have provided significant environmental benefits^29,65^ and opened new opportunities across the broad fields of biomedical data science^22–28^. The novel pseudo-training methods we developed provide several advantages for cloud-based PRS model training solutions. First, pseudo-training mitigates the privacy risks and concerns associated with uploading or sharing individual-level genetic datasets online. Second, individual-level validation data is not always available for PRS model development, and our pseudo-training pipeline provides a more accessible solution for PRS training across a wider range of disease and health outcomes. Third, pseudo-training could deliver PRS with better prediction performance, especially for those disease outcomes with limited individual-level tuning data available (e.g., *N_tuning_* < 1,000). The pseudo-training approach we developed would lend power for these understudied outcomes. Fourth, the pseudo-training approach facilitates seamless integration with online GWAS data resources, such as the GWAS Catalog, providing a centralized data resource for PRS model training. In the future, we plan to extend the functionality of pseudo-training and PennPRS in several directions. For example, the current training procedure relies on the five super ancestral population labels^34^ (e.g., EUR and AFR). We aim to expand our framework to include additional population labels and further integrate flexible genetic ancestry continuum information^66^ as the field increasingly incorporates diverse and admixed ancestry information^18^. We will also provide unified PRS models for the general population instead of ancestry-specific PRS models that require categorizing individuals into discrete ancestry groups^15^. Furthermore, beyond generating PRS model training, we will additionally develop pseudo-training methods to produce additional accuracy metrics and uncertainty measures for the generated PRS models, such as confidence intervals^67^, which are increasingly critical for downstream clinical applications of PRS^68–71^.

Notably, the applicability of the FAIR data principles^72^ to our cloud computing platform highlights the broad impact of PennPRS on future translational research involving PRS. By providing standardized computing pipelines, curated data resources, detailed documentation, and accessible PRS weight files, PennPRS facilitates transparency and ensures the **F**indability, **A**ccessibility, **I**nteroperability, and **R**eusability of PRS resources. This is particularly important as the adoption and application of PRS continue to expand in precision medicine, a process that typically involves multiple steps, from PRS model development and assessment to implementation and translation in clinical settings. Efficient data and information sharing between these stages will be critical for successful translation. In addition, the computing methods and data resources provided by our PennPRS platform enable efficient between study comparisons of PRS within the same ancestry background. This is particularly important given the diversity of biobanks in the US and globally. By providing a centralized platform, PennPRS anchors comparisons to a consistent linkage disequilibrium and allele frequency background, reducing bias or noise that might hinder cross-study comparisons. This allows researchers to focus on other factors affecting performance^18,73^, improving the reliability and generalizability of PRS analyses across studies.

There are also limitations to cloud computing platforms. For example, if users have already established powerful local computational clusters—typically supported by their research institutions’ infrastructure—they might consider setting up the pipeline locally, although this approach may be more time consuming and less environmentally friendly^65^. To meet this complementary need, we have developed a ready-to-use offline version of PennPRS pipeline. Another challenge of cloud computing platform is the maintenance of the server and pipelines. To ensure sustainable development of our platform, we have formed a dedicated interdisciplinary team of researchers centered around the PRS research community at the University of Pennsylvania, in collaboration with researchers from other institutions, to support regular updates to PennPRS. These updates will include incorporating additional PRS methods, generating new data resources, and more efficient method implementations. The long-term maintenance of the platform is supported by the robust AWS server, with additional technical assistance from our local IT teams. We welcome user feedback and suggestions to improve the PennPRS platform and better meet the diverse needs of the global PRS research community.

## Acknowledgements

Research reported in this publication was supported by the National Human Genome Research Institute under Award Number 5R00HG012223 (J.J.), National Institute of Mental Health under Award Number R01MH136055 (B.Z.), and National Institute on Aging under Award Number RF1AG082938 (B.Z.). The content is solely the responsibility of the authors and does not necessarily represent the official views of the National Institutes of Health. The study has also been partially supported by funding from the Purdue University Statistics Department, Department of Statistics and Data Science at the University of Pennsylvania, Wharton Dean’s Research Fund, Analytics at Wharton, Wharton AI & Analytics Initiative, Perelman School of Medicine CCEB Innovation Center Grant, and the University Research Foundation at the University of Pennsylvania. The individual-level genotype and phenotype data for UK Biobank validation samples used in this study were obtained under application 76139 subject to a data transfer agreement. We would like to thank the individuals who represented themselves in the UK Biobank for their participation and the research teams for their efforts in collecting, processing, and disseminating these datasets. We would like to thank the research computing and IT groups at the Wharton School of the University of Pennsylvania and the Rosen Center for Advanced Computing at the Purdue University for providing computational resources and support that have contributed to these research results.

## Author Contributions Statement

J.J. and B.Z. conceived the project. J.J. developed the pseudo-training methods and algorithms. B.L. and X.W. set up the cloud computing server with the help from local IT teams. J.J., B.L., X.W., and B.Z. carried out data analyses, interpreted the results, designed the cloud computing platform, and developed the offline pipeline. X.Y., Y.L., J.S., R.W., and Z.F. processed the GWAS summary statistics, developed the curated datasets, and contributed to the development and testing of the computing platform and offline pipeline. C.Y., F.X., T.G., M.D.R., G.W., B.P., and R.W. contributed to the design of the methods, cloud computing platform, and offline pipeline. J.J. and B.Z. drafted the manuscript with feedback from all authors.

## Methods

### Single-ancestry PRS pseudo-training

Single-ancestry PRS training aims to develop PRS models for a target genetic ancestral population based on a single GWAS summary dataset generated from training samples of the same population. We developed a general summary data-based parameter optimization approach for multiple single-ancestry PRS methods that avoids the need for individual-level tuning data (**Supplementary Fig. 1a**). We have implemented the approach to develop pseudo-training versions of three single-ancestry PRS methods: C+T-pseudo, LDpred2-pseudo, and Lassosum2-pseudo, which are included in our PennPRS cloud computing platform. We have also developed the pseudo-training version of an additional method, PRS-CS-grid-pseudo, which has a much higher computational demand and is included in our offline pipeline. Our PRS pseudo-training pipeline follows the general framework of PUMAS^31,33^. Specifically, in Step 1, we use the subsampling approach in PUMAS to sample marginal association statistics for two “pseudo” subsets of training and validation individuals from the full GWAS summary data^31^. This approach enables us to generate GWAS summary statistics for pseudo training and validation sets for PRS training and parameter tuning, respectively, without the need to collect an independent individual-level dataset for parameter tuning. In Step 2, we apply each selected PRS method to train PRS models on the pseudo summary-level training dataset. In Step 3, we conduct parameter tuning on the pseudo summary-level validation dataset. This summary data-based parameter tuning is conducted using the method in PUMAS that allows estimation of the prediction *R*^2^ of PRS using summary statistics only. This step selects the best tuning parameter setting for each method based on performance on the pseudo validation summary dataset. If multiple PRS methods are implemented in Step 2, we will proceed to Step 4, which offers the option to train ensemble PRS models. These models combine the PRS models generated by various methods using the pseudo-validation dataset and two ensemble approaches: Ensemble-pseudo and Ensemble-ARM-pseudo, which will be introduced in the next section. Finally, in Step 5, we train the final PRS models on the full GWAS summary dataset with selected tuning parameter settings obtained from Step 3 and trained ensemble weights for different methods in the ensemble PRS models from Step 4. To increase stability of the parameter tuning results, we repeat the training-validation data splitting procedure in Step 2 k=2 times and conduct Steps 2 to 4 with k-fold cross-validation. Specifically, for parameter tuning, we select the parameter setting that correspond to the highest estimated prediction *R*^2^ on the pseudo validation data averaged across the k folds; and for ensemble PRS training, we obtain the weights in the ensemble model as the average across the k folds.

We have identified several potential issues of the original PUMAS algorithm when incorporating it with different PRS methods and have made substantial modifications accordingly to ensure the applicability and increase the robustness of our pipeline. For example, the original Lassosum2 and LDpred2 algorithms may generate non-convergent or problematic PRS weights (e.g., overly large 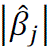) under some tuning parameter settings, which can lead to inflated *R*^2^ estimate for these settings, resulting in problematic parameter tuning by PUMAS. We resolved this issue by discarding the tuning parameter settings in which there exist genetic effect estimates 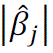 > 1. Furthermore, it is likely that the final PRS model trained on the full GWAS summary data has non-convergent variant weights, even though the selected optimal tuning parameter setting gives a converged model when trained on the pseudo training dataset. This issue is due to the unstable performance of the PRS methods, not the PUMAS pseudo-training algorithm itself. To avoid this inconsistency in the PRS models trained based on the pseudo-training dataset and the original summary dataset, we select optimal tuning parameters only from the settings that lead to converged variant weights on the original summary data. We also noticed that the selected tuning parameter setting may be far from the optimal setting if its adjacent tuning parameter settings led to nonconvergent results. We thus only consider parameter settings for which the adjacent settings also lead to converged results. If no such candidate setting exists, then we will skip this step and just select the setting that gives the highest *R*^2^ on the pseudo validation set. Finally, for traits that have minimal heritability or have a small GWAS sample size, the PRS model trained by some of the methods may have limited power, reflected by negative prediction *R*^2^ estimates on the pseudo validation data. In this case, we still output the trained PRS models but will also print a warning message to let users know about this issue. We will also exclude the corresponding PRS models from the pseudo ensemble learning step.

### Pseudo ensemble learning combining multiple single-ancestry PRS models

As mentioned in the previous section, if multiple PRS methods are implemented in single-ancestry analysis, we will provide an option to conduct pseudo-training of ensemble PRS models combining PRS trained by the various methods based on the pseudo validation dataset. We propose two approaches for the pseudo ensemble PRS training. The first approach trains a linear combination^39^ of the PRS models obtained from the various methods (“Ensemble-pseudo”). This approach was proposed in the PUMA-CUBS framework^33^. We notice that this approach sometimes generates a PRS that has a lower power than the best single PRS model, possibly due to the suboptimal performance of some of the single PRS models. Therefore, we propose an alternative approach adopting a model combination method named adaptive regression by mixing (ARM)^40^, which, under certain conditions, can approximate the optimal performance among a set of single models (“Ensemble-ARM-pseudo”). We observe from our simulation studies and data applications that either one of the two approaches outperforms the other on different phenotypes and with different training GWAS datasets. We thus provide both ensemble PRS models to users to further increase the power of the “best” PRS model provided by PennPRS.

### Multi-ancestry PRS pseudo-training

Multi-ancestry PRS training jointly analyzes ancestry-stratified GWAS summary statistics from *K* ancestral populations (a subset of [EUR, AFR, AMR, EAS, and SAS]) and generates ancestry-specific PRS models for the *K* populations. We developed a GWAS summary data-based parameter tuning approach for multi-ancestry PRS training that avoids the need for individual-level tuning data (**Supplementary Fig. 1b**). We have implemented this approach to develop the pseudo-training version of PROSPER on our PennPRS cloud computing platform and two other methods, PRS-CSx-pseudo and MUSSEL-pseudo, which require much larger memory and/or are computationally more intensive and are thus only included in our offline pipeline.

Our general multi-ancestry PRS pseudo-training framework follows that of PUMAS^31,33^. Specifically, in Step 1, we use the subsampling approach in PUMAS to generate summary statistics for pseudo training and validation sets for PRS training and parameter tuning, respectively, for each of the *K* ancestry populations. In Step 2, we apply each selected method to train PRS models on the pseudo training dataset. For PROSPER-pseudo and MUSSEL-pseudo which prerequire implementation of the single-ancestry Lassosum2 and LDpred2 algorithms, respectively, we use a procedure similar to the one in single-ancestry pseudo-training for selecting optimal parameters of Lassosum2-pseudo and LDpred2-pseudo. In Step 3, we conduct parameter optimization of the multi-ancestry joint modeling step in PROSPER or MUSSEL on the pseudo summary-level validation dataset. This step selects a best PRS model for each ancestry based on its performance (the estimated prediction *R*^2^) on the pseudo validation dataset. All three methods have a final ensemble learning step (Step 4, **Supplementary Fig. 1**), where PRS-CSx trains a linear combination of the *K* best ancestry-specific PRS models from Step 3, while PROSPER and MUSSEL use the super learner algorithm^74^ with base learners including linear regression, elastic-net regression, and ridge regression to train an “optimal” linear combination of all PRS models across all tuning parameter settings and ancestries. For PRS-CSx, we use the ensemble approach in PUMAS to train the final PRS model for each ancestry based on the pseudo validation dataset of that ancestry. For PROSPER and MUSSEL, a summary data version of the super learner can be implemented for the final ensemble step utilizing a recently developed approach^75^. For now, we consider an alternative strategy, where we train a linear combination of a subset of *L* best performing single PRS models without regularization. Specifically, for each ancestry, we first select the top *L* of the 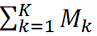 PRS models that have the highest prediction *R*^2^ on the pseudo validation dataset of that ancestry, where *M*” denotes the number of tuning parameter settings, i.e., number of candidate PRS models, generated for ancestry *k*, *k* = 1,2, …, *K*. We then train a linear combination of these *L* top PRS models on the pseudo validation dataset. We set *L* to the minimum between five and the number of converged PRS models among the 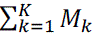 models. Finally, in Step 5, we train the final PRS models on the full original GWAS summary data based on the selected optimal parameter settings from Step 3 and trained ensemble weights for different single PRS models from Step 4. We notice that the single best PRS model may have a higher prediction power than the final ensemble PRS. We thus provide both the best single PRS model (“PROSPER-Single-pseudo PRS”, **Figs. 1c** and **5**, **Supplementary Figs. 4-5**) and the final PRS model with the ensemble step (“PROSPER-pseudo PRS”, **Figs. 1c** and **5, Supplementary Figs. 4-5**) to the user. Again, we repeat the training-validation data splitting procedure in Step 2 k=2 times and conduct Steps 2-4 with k-fold cross-validation to increase stability of the results. Specifically, for parameter tuning, we select the parameter setting that correspond to the highest estimated prediction *R*^2^ on the pseudo validation data averaged across the k folds; and for the ensemble step, we obtain the weights in the ensemble model as the average across the k folds. In our multi-ancestry analysis pipeline, we also consider the various modifications to the original PUMAS algorithm implemented in our single-ancestry analysis pipeline to improve robustness of the pseudo-training.

### Configuration of the online PennPRS pipeline

Our PennPRS cloud computing pipeline currently supports seven PRS methods, including pseudo-training versions of three single-ancestry methods, C+T-pseudo, lassosum2-pseudo, and LDpred2-pseudo; three tuning-parameter-free single-ancestry methods, PRS-CS-auto, LDpred2-auto, and DBSLMM; and pseudo-training version of one multi-ancestry method, PROSPER-pseudo. PennPRS supports PRS model development based on genetic variants in the HapMap 3^76^. For implementation of methods that have tuning parameters, we set up default tuning parameter settings based on the ones in the original algorithms of the methods but with slight modifications to balance between prediction performance and computational efficiency of our online PRS training. We use genotype data of unrelated individuals from the Phase 3 1000 Genomes project as the linkage disequilibrium (LD) reference data. We now introduce the parameter settings and other relevant details of the newly developed pseudo-training methods supported by PennPRS.

#### C+T-pseudo

C+T-pseudo first conducts an LD clumping step to select relatively independent genetic variants with an absolute pairwise correlation lower than *r*^2^ = 0.1 within a genetic distance 500kb calculated based on the reference genotype dataset for the same ancestral population from the Phase 3 1000 Genomes Project^34^. It then selects the remaining genetic variants that reach varying p-value cutoffs (tuning parameter: pt)^4,5^. Our default candidate values for pt are 5×10^−8^, 5×10^−7^, 5×10^−6^, 5×10^−5^, 5×10^−4^, 5×10^−3^, 5×10^−2^, and 5×10^−1^. C+T-pseudo then selects the score with the “optimal” p-value threshold based on the performance on the pseudo validation dataset with respect to the prediction *R*^2^. PennPRS runs C+T-pseudo using PLINK 1.90^77^.

#### Lassosum2-pseudo

Lassosum2-pseudo is a penalized regression-based approach that estimates joint genetic effect sizes based on GWAS summary statistics. Tuning parameters include (i) λ: shrinkage parameter in the *L*_2_ regularization (default candidate values: 0.001, 0.01, 0.1, and 1); (ii) number of candidate values for λ, the shrinkage parameter in the *L*_1_ regularization (default: 30); and (iii) ratio between the lowest and highest candidate values of λ (default: 0.01). The current version of PennPRS implements Lassosum2-pseudo with R package “bigsnpr” (version 1.6.1, last updated Jun 8, 2023).

#### LDpred2-pseudo

LDpred2-pseudo is a Bayesian approach that jointly analyzes correlated genetic variants across the genome and accounts for LD^37,38^. It uses a spike-and-slab prior on genetic effect sizes, assuming a proportion (*p*) of the genetic variants have non-zero effect on the phenotype. Tuning parameters include (1) the causal variant proportion *p* (default candidate values: 10^−5^, 3.2×10^-5^, 10^-4^, 3.2 × 10^-4^, 10^-3^, 3.2 × 10^-3^, 10^-2^, 3.2 × 10^-2^, 10^-1^, 3.2 × 10^-1^, and 1), (2) heritability-related parameter, α: the total heritability is set to 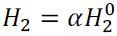, where 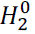 is the heritability estimated by LD score regression^78^ (default candidate values: α = 0.7, 1.0, and 1.4), and an additional sparse option (default: FALSE) to shrink the posterior genetic effects that exceed *p* to zero. The current version of PennPRS implements LDpred2-pseudo with R package “bigsnpr” (version 1.6.1, last updated Jun 8, 2023).

#### PROSPER-pseudo

PROSPER-pseudo is a penalized regression-based multi-ancestry PRS method that utilizes an *L*_1_ penalty to induce sparsity of genetic variants with non-zero effects and an *L*_2_ penalty to induce correlation in genetic effects between ancestries^41^. Tuning parameters include (i) the number of candidate values of the shrinkage parameter in the *L*_1_ penalty (default: 5) and (ii) the number of candidate values of the shrinkage parameter in the *L*_2_ penalty (default: 5).

### Simulation studies

We evaluated the performance of our proposed pseudo-training approach for both single– and multi-ancestry PRS development in comparison to the traditional, individual-level tuning data-based PRS training in various data settings based on a large-scale synthetic GWAS data previously generated^42^. The synthetic genotype data were generated for all five super populations (EUR, AFR, AMR, EAS, and SAS) using HAPGEN2 (version 2.1.2)^79^ to closely mimic the reference genotype data from the Phase 3 1000 Genomes Project. Phenotype data were generated assuming a causal variant proportion of 1%, 0.1%, of 0.05% and GWAS sample size of 15,000, 45,000, or 80,000 across the five ancestral populations. Details of the simulation procedure were previously described^42^.

### Real data analyses for evaluation of pseudo PRS training

#### UKB imaging data analysis

We conducted a large-scale evaluation of single-ancestry pseudo-training methods using multi-organ multi-modality imaging data^48,80,81^ from the UK Biobank (UKB) study, covering brain, heart, eye, and abdominal organs (**Supplementary Tables 2-5**). For the brain, we used imaging-derived phenotypes from three major modalities: structural MRI (sMRI), diffusion MRI (dMRI), and resting-state functional MRI (rfMRI). For example, brain sMRI included 1,432 phenotypes generated from the FIRST, FAST, and FreeSurfer pipelines^44^. Brain dMRI data included 674 phenotypes processed with TBSS and ProbtrackX, while brain rfMRI encompassed 82 phenotypes from whole-brain spatial independent component analysis^44,81–83^, covering regional amplitude and global functional connectivity. For cardiac MRI, we used 82 phenotypes related to the heart and aorta^46,84^. Additionally, 41 abdominal MRI phenotypes^85–94^ were included, covering kidney, liver, and abdominal organ or tissues. In addition, we analyzed 46 phenotypes derived from eye optical coherence tomography images^47^. The GWAS summary data for these imaging phenotypes were obtained from subjects of self-reported British European ancestry, with average sample sizes of 32,634 for brain, 30,506 for heart, 29,849 for abdomen, and 50,465 for eye. Age, sex, the top 40 genetic principal components (PCs), and imaging-specific covariates were adjusted for, as detailed in a previous study^45,48^. PRS performance was assessed on 2,227 to 8,172 European non-British subjects, using the same set of covariates as those in the corresponding GWAS.

#### FinnGen disease data analysis

We evaluated the performance of single-ancestry pseudo-training methods on binary phenotypes using GWAS summary statistics from the FinnGen study (R9)^49^. Following the FinnGen-phecode mapping approach used in previous studies^48,50^, we mapped 29 disease pairs, with an average of 333,355 cases and controls per phenotype (**Supplementary Tables 6-7**). PRS performance was assessed on 1,225 to 155,170 European cases from the UKB, with adjustments for effects of age and sex.

#### UKB-PPP Olink plasma protein data analysis

We evaluated the performance of single-ancestry pseudo PRS training methods on 2,734 Olink plasma proteins from the UKB-PPP^51^ project (**Supplementary Tables 8**-**9**). The GWAS summary statistics were obtained from a previous study^95^, which included 40,852 subjects of British European ancestry with adjustments for age, sex, and the top 40 genetic PCs. PRS performance was assessed on 2,517 to 2,923 European non-British subjects, using the same set of covariates as in the GWAS.

#### GLGC blood lipids data analysis

We trained ancestry-specific PRS models by both pseudo-training and individual-level tuning versions of the various methods for four blood lipids, including high-density lipoprotein (HDL), low-density lipoprotein (LDL), log-transformed triglycerides (logTG), and total cholesterol (TC)^54^. The ancestry-stratified training GWAS summary data were obtained from the Global Lipids Genetics Consortium^54^ (GLGC) on five ancestry groups including EUR (*N* = 840,018-927,975), AFR or admixed AFR (*N* = 87,759-92,554), Hispanic/Latino (*N* = 33,989-48,056), EAS (*N* = 80,676-145,512), and SAS (*N* = 33,658-34,135)^54^.

We validated method performance on a random set of 20,000 UKB individuals of EUR ancestry and all UKB individuals of AFR (*N* = 9,169), AMR (N = 750), EAS (*N* = 2,019), and SAS (*N* = 10,853) ancestry. We inferred the ancestry of the UKB individuals by a genetic component analysis^41^. We used 50% of these UKB samples to conduct individual-level data-based parameter tuning, ensemble PRS training, and conducting the ensemble step in PROSPER, and used the remaining 50% (testing set) to evaluate PRS performance of the various methods. GWAS sample sizes, validation sample sizes, and the number of genetic variants analyzed are reported in **Supplementary Table 11**. Detailed data quality control procedures were previously described^42,55^. Age, gender, and the top 10 genetic PCs were adjusted for as covariates when calculating prediction *R*^2^ of the PRS models.

#### UKB/CHIMGEN brain imaging data analysis

We evaluated the performance of multi-ancestry pseudo-training methods on brain imaging phenotypes using GWAS summary statistics from both the Chinese Imaging Genetics (CHIMGEN) study^56^ for East Asians (average *N* = 7,058) and the UKB study for British European ancestry (average *N* = 34,286). Similar to our single-ancestry analysis on UKB, we included 968 phenotypes from sMRI and 445 from dMRI. PRS performance was assessed on 443 Asian subjects from the UKB study (half were used as tuning samples for parameter optimization for traditional PRS training methods, half were used as testing data to evaluate performance of all methods) with adjustments for the same set of covariates as in the single-ancestry analysis on the same phenotypes. GWAS sample sizes, validation sample sizes, and the number of genetic variants analyzed are reported in **Supplementary Table 13**.

### Other GWAS datasets on which we have generated PRS models

We applied our offline pipeline to GWAS summary statistics from the Biobank Japan (BBJ)^62^, the Million Veteran Program (MVP) study^63^, the Global Biobank Meta-analysis Initiative (GBMI) consortium^64^, and the GWAS Catalog^57^. For the BBJ, we conducted single-ancestry analysis on GWAS summary statistics for 169 phenotypes available at https://pheweb.jp/downloads. For the GWAS Catalog, we analyzed nearly 8,000 harmonized datasets in single-ancestry analysis. For the GBMI, we performed multi-ancestry analysis on nine phenotypes with ancestry-stratified GWAS summary statistics from the five super populations^34^. For the MVP study, we carried out multi-ancestry analysis on 181 phenotypes with ancestry-stratified GWAS summary statistics from AFR, AMR, EAS, and EUR populations. Using default parameter settings, our pipelines were successfully applied to these data resources, and the generated PRS models have been shared in the PennPRS public resource hub.

### Cloud computing platform development

PennPRS is a cloud-based platform hosted on AWS that consists of two main components: the frontend and the backend. For the frontend, we used Next.js (https://nextjs.org/) and MUI (https://mui.com/) to create a clean, intuitive interface. Users can easily input their data (through file uploads or data queries), choose the type of analysis, PRS methods, and parameter setting they want, and view job status and results. We used Next.js to ensure that the platform loads quickly and that all interactions (such as submitting data or viewing outputs) are smooth and responsive. The backend provides the infrastructure for PRS model development, including data harmonization and QC pipelines, GWAS Catalog data querying, and various PRS methods and training mechanisms. We developed the backend with FastAPI (https://fastapi.tiangolo.com/), which allows us to process multiple tasks efficiently, supporting both simple requests and more complex data processing. For example, when a user uploads data for PRS analysis, FastAPI sends this data to the job queue, ensuring that requests are processed in a fair and timely manner.

In addition, Redis (https://redis.io/) is used for job management and queue, keeping track of all incoming requests and organizing them so that the system can handle multiple tasks simultaneously. Redis also helps prevent delays and keeps the platform running smoothly even during busy times. Moreover, since different types of analyses have different resource requirements, we organized the computing infrastructure into distinct subgroups to optimize resources. Each subgroup is tailored to handle specific types of jobs, ensuring that the right resources (such as memory and CPU power) are available for the task at hand, which optimizes resource allocation and improves overall efficiency. Once the analysis is completed, the results are sent back to the frontend so users can access and download them. To ensure reliability and scalability, the platform incorporates monitoring tools for system performance checks, automated testing, and continuous integration pipelines. This setup enables quick future updates and secure data handling, ensuring a smooth user experience as demand grows.

## Code availability

The developed PRS pseudo-training methods and PennPRS pipelines can be freely accessed at https://pennprs.org/ and https://github.com/PennPRS/pipeline.

## Data availability

The simulated genotype and phenotype data used in our simulations are available at: https://dataverse.harvard.edu/dataset.xhtml?persistentId=doi:10.7910/DVN/COXHAP. GWAS summary statistics used in our PRS training and evaluation can be obtained from their respective data sources, subject to data sharing policies and approvals. Specifically, the harmonized GWAS summary statistics from the GWAS Catalog are available at https://www.ebi.ac.uk/gwas/downloads/summary-statistics. The EUR GWAS summary statistics for the UKB imaging phenotypes across different organs are available from previous study^45,48^. The EUR protein GWAS summary statistics from the UKB-PPP project are available from previous study^95^. The EUR GWAS summary statistics from the FinnGen study are available at https://www.finngen.fi/en/access_results. The EAS GWAS summary statistics from BBJ are available at https://pheweb.jp/. The EAS GWAS summary statistics for brain imaging phenotypes from the CHIMGEN study are available from previous study^56^. Ancestry-stratified GWAS summary statistics from the GBMI are available at https://www.globalbiobankmeta.org/resources. Ancestry-stratified GWAS summary statistics for blood lipids across five super populations from GLGC are available at http://csg.sph.umich.edu/willer/public/glgc-lipids2021/results/ancestry_specific. Ancestry-stratified GWAS summary statistics from the MVP study are available from previous study^63^. The individual-level UK Biobank data used in this study can be requested from https://www.ukbiobank.ac.uk/. The PRS model weights generated by the PennPRS pipeline have been made publicly available through the PennPRS public resource hub at https://pennprs.org/result.

## References

1. Uffelmann, E. et al. Genome-wide association studies. Nature Reviews Methods Primers 1, 1–21 (2021).

2. Abdellaoui, A., Yengo, L., Verweij, K.J. & Visscher, P.M. 15 years of GWAS discovery: Realizing the promise. The American Journal of Human Genetics (2023).

3. Choi, S.W., Mak, T.S.-H. & O’Reilly, P.F. Tutorial: a guide to performing polygenic risk score analyses. Nature protocols 15, 2759–2772 (2020).

4. Wray, N.R., Goddard, M.E. & Visscher, P.M. Prediction of individual genetic risk to disease from genome-wide association studies. Genome research 17, 1520–1528 (2007).

5. Consortium, I.S. Common polygenic variation contributes to risk of schizophrenia and bipolar disorder. Nature 460, 748–752 (2009).

6. Torkamani, A., Wineinger, N.E. & Topol, E.J. The personal and clinical utility of polygenic risk scores. Nature Reviews Genetics 19, 581–590 (2018).

7. Kullo, I.J. et al. Polygenic scores in biomedical research. Nature Reviews Genetics 23, 524–532 (2022).

8. Chatterjee, N., Shi, J. & García-Closas, M. Developing and evaluating polygenic risk prediction models for stratified disease prevention. Nature Reviews Genetics 17, 392 (2016).

9. Lewis, C.M. & Vassos, E. Polygenic risk scores: from research tools to clinical instruments. Genome medicine 12, 1–11 (2020).

10. Wand, H. et al. Improving reporting standards for polygenic scores in risk prediction studies. Nature 591, 211–219 (2021).

11. Ma, Y. & Zhou, X. Genetic prediction of complex traits with polygenic scores: a statistical review. Trends in Genetics 37, 995–1011 (2021).

12. Yang, S. & Zhou, X. PGS-server: accuracy, robustness and transferability of polygenic score methods for biobank scale studies. Briefings in Bioinformatics 23, bbac039 (2022).

13. Lambert, S.A. et al. The Polygenic Score Catalog as an open database for reproducibility and systematic evaluation. Nature Genetics 53, 420–425 (2021).

14. Page, M.L. et al. The Polygenic Risk Score Knowledge Base offers a centralized online repository for calculating and contextualizing polygenic risk scores. Commun Biol 5, 899 (2022).

15. Kachuri, L. et al. Principles and methods for transferring polygenic risk scores across global populations. Nature Reviews Genetics, 1–18 (2023).

16. Duncan, L. et al. Analysis of polygenic risk score usage and performance in diverse human populations. Nature communications 10, 1–9 (2019).

17. Hou, K. et al. Admix-kit: an integrated toolkit and pipeline for genetic analyses of admixed populations. Bioinformatics 40, btae148 (2024).

18. Ding, Y. et al. Polygenic scoring accuracy varies across the genetic ancestry continuum. Nature 618, 774–781 (2023).

19. Kullo, I.J. et al. The PRIMED Consortium: Reducing disparities in polygenic risk assessment. The American Journal of Human Genetics 111, 2594–2606 (2024).

20. Linder, J.E. et al. Returning integrated genomic risk and clinical recommendations: The eMERGE study. Genetics in Medicine 25, 100006 (2023).

21. Lennon, N.J. et al. Selection, optimization and validation of ten chronic disease polygenic risk scores for clinical implementation in diverse US populations. Nature Medicine 30, 480–487 (2024).

22. Watanabe, K., Taskesen, E., Bochoven, A. & Posthuma, D. Functional mapping and annotation of genetic associations with FUMA. Nature Communications 8, 1826 (2017).

23. Das, S. et al. Next-generation genotype imputation service and methods. Nature genetics 48, 1284–1287 (2016).

24. Li, Y. et al. Analyzing bivariate cross-trait genetic architecture in GWAS summary statistics with the BIGA cloud computing platform. bioRxiv, 2023.04.28.538585 (2023).

25. Dai, C. et al. quantms: a cloud-based pipeline for quantitative proteomics enables the reanalysis of public proteomics data. Nature Methods, 1–5 (2024).

26. Hayashi, S. et al. brainlife. io: a decentralized and open-source cloud platform to support neuroscience research. Nature methods 21, 809–813 (2024).

27. Artomov, M., Loboda, A.A., Artyomov, M.N. & Daly, M.J. Public platform with 39,472 exome control samples enables association studies without genotype sharing. Nature Genetics 56, 327–335 (2024).

28. Langmead, B. & Nellore, A. Cloud computing for genomic data analysis and collaboration. Nature Reviews Genetics 19, 208–219 (2018).

29. Lannelongue, L., Grealey, J. & Inouye, M. Green Algorithms: Quantifying the Carbon Footprint of Computation. Adv Sci (Weinh) 8, 2100707 (2021).

30. Chen, T., Zhang, H., Mazumder, R. & Lin, X. Fast and scalable ensemble learning method for versatile polygenic risk prediction. Proceedings of the National Academy of Sciences 121, e2403210121 (2024).

31. Zhao, Z. et al. PUMAS: fine-tuning polygenic risk scores with GWAS summary statistics. Genome biology 22, 1–19 (2021).

32. Zhang, Q., Privé, F., Vilhjálmsson, B. & Speed, D. Improved genetic prediction of complex traits from individual-level data or summary statistics. Nature communications 12, 4192 (2021).

33. Zhao, Z. et al. Optimizing and benchmarking polygenic risk scores with GWAS summary statistics. Genome Biol 25, 260 (2024).

34. Genomes Project, C., et al. A global reference for human genetic variation. Nature 526, 68–74 (2015).

35. Mak, T.S.H., Porsch, R.M., Choi, S.W., Zhou, X. & Sham, P.C. Polygenic scores via penalized regression on summary statistics. Genet Epidemiol 41, 469–480 (2017).

36. Privé, F., Arbel, J., Aschard, H. & Vilhjálmsson, B.J. Identifying and correcting for misspecifications in GWAS summary statistics and polygenic scores. Human Genetics and Genomics Advances 3(2022).

37. Privé, F., Arbel, J. & Vilhjálmsson, B.J. LDpred2: better, faster, stronger. Bioinformatics 36, 5424–5431 (2020).

38. Vilhjalmsson, B.J. et al. Modeling Linkage Disequilibrium Increases Accuracy of Polygenic Risk Scores. Am J Hum Genet 97, 576–92 (2015).

39. Prive, F., Vilhjalmsson, B.J., Aschard, H. & Blum, M.G.B. Making the Most of Clumping and Thresholding for Polygenic Scores. Am J Hum Genet 105, 1213–1221 (2019).

40. Yang, Y. Adaptive Regression by Mixing. Journal of the American Statistical Association 96, 574–588 (2001).

41. Zhang, J. et al. An ensemble penalized regression method for multi-ancestry polygenic risk prediction. Nature Communications 15, 3238 (2024).

42. Zhang, H. et al. A new method for multiancestry polygenic prediction improves performance across diverse populations. Nat Genet 55, 1757–1768 (2023).

43. Littlejohns, T.J. et al. The UK Biobank imaging enhancement of 100,000 participants: rationale, data collection, management and future directions. Nature communications 11, 1–12 (2020).

44. Alfaro-Almagro, F. et al. Image processing and Quality Control for the first 10,000 brain imaging datasets from UK Biobank. NeuroImage 166, 400–424 (2018).

45. Fan, Z. et al. The role of sleep in the human brain and body: insights from multi-organ imaging genetics. medRxiv, 2022.09. 08.22279719 (2022).

46. Bai, W. et al. A population-based phenome-wide association study of cardiac and aortic structure and function. Nature Medicine 26, 1654–1662 (2020).

47. Zhao, B. et al. Eye-brain connections revealed by multimodal retinal and brain imaging genetics. Nature Communications 15, 6064 (2024).

48. Yang, X. et al. Multi-organ imaging-derived polygenic indexes for brain and body health. medRxiv, 2023.04. 18.23288769 (2023).

49. Kurki, M.I. et al. FinnGen provides genetic insights from a well-phenotyped isolated population. Nature 613, 508–518 (2023).

50. Sun, B.B. et al. Genetic associations of protein-coding variants in human disease. Nature 603, 95–102 (2022).

51. Sun, B.B. et al. Plasma proteomic associations with genetics and health in the UK Biobank. Nature 622, 329–338 (2023).

52. Deng, Y.-T. et al. Atlas of the plasma proteome in health and disease in 53,026 adults. Cell 188, 253–271. e7 (2025).

53. Xu, Y. et al. An atlas of genetic scores to predict multi-omic traits. Nature 616, 123–131 (2023).

54. Graham, S.E. et al. The power of genetic diversity in genome-wide association studies of lipids. Nature 600, 675–679 (2021).

55. Jin, J., et al. MUSSEL: Enhanced Bayesian Polygenic Risk Prediction Leveraging Information across Multiple Ancestry Groups. bioRxiv (2023).

56. Fu, J. et al. Cross-ancestry genome-wide association studies of brain imaging phenotypes. Nature Genetics, 1–11 (2024).

57. Sollis, E. et al. The NHGRI-EBI GWAS Catalog: knowledgebase and deposition resource. Nucleic Acids Res 51, D977–D985 (2023).

58. Ge, T., Chen, C.-Y., Ni, Y., Feng, Y.-C.A. & Smoller, J.W. Polygenic prediction via Bayesian regression and continuous shrinkage priors. Nature Communications 10, 1776 (2019).

59. Yang, S. & Zhou, X. Accurate and scalable construction of polygenic scores in large biobank data sets. The American Journal of Human Genetics 106, 679–693 (2020).

60. Fernández-Rhodes, L. et al. Ancestral diversity improves discovery and fine-mapping of genetic loci for anthropometric traits—The Hispanic/Latino Anthropometry Consortium. Human Genetics and Genomics Advances 3(2022).

61. Ruan, Y. et al. Improving polygenic prediction in ancestrally diverse populations. Nat Genet 54, 573–580 (2022).

62. Sakaue, S. et al. A cross-population atlas of genetic associations for 220 human phenotypes. Nature genetics 53, 1415–1424 (2021).

63. Verma, A. et al. Diversity and scale: Genetic architecture of 2068 traits in the VA Million Veteran Program. Science 385, eadj1182 (2024).

64. Zhou, W. et al. Global Biobank Meta-analysis Initiative: Powering genetic discovery across human disease. Cell Genomics 2, 100192 (2022).

65. Lannelongue, L. et al. GREENER principles for environmentally sustainable computational science. Nature Computational Science 3, 514–521 (2023).

66. Ruan, Y. et al. Leveraging genetic ancestry continuum information to interpolate PRS for admixed populations. medRxiv, 2024.11.09.24316996 (2024).

67. Ding, Y. et al. Large uncertainty in individual polygenic risk score estimation impacts PRS-based risk stratification. Nature genetics 54, 30–39 (2022).

68. Abramowitz, S.A. et al. Evaluating Performance and Agreement of Coronary Heart Disease Polygenic Risk Scores. JAMA (2024).

69. Momin, M.M., Lee, S., Wray, N.R. & Lee, S.H. Significance tests for R2 of out-of-sample prediction using polygenic scores. The American Journal of Human Genetics 110, 349–358 (2023).

70. Wang, X. et al. Impact of individual level uncertainty of lung cancer polygenic risk score (PRS) on risk stratification. Genome medicine 16, 22 (2024).

71. Fu, H., Huang, J., Fan, Z. & Zhao, B. Uncertainty of high-dimensional genetic data prediction with polygenic risk scores. arXiv preprint arXiv:2412.20611 (2024).

72. Wilkinson, M.D. et al. The FAIR Guiding Principles for scientific data management and stewardship. Scientific data 3, 1–9 (2016).

73. Wang, Y. et al. Theoretical and empirical quantification of the accuracy of polygenic scores in ancestry divergent populations. Nature communications 11, 1–9 (2020).

74. van der Laan, M.J., Polley, E.C. & Hubbard, A.E. Super learner. Stat Appl Genet Mol Biol 6, Article25 (2007).

75. Zhao, Z. et al. One score to rule them all: regularized ensemble polygenic risk prediction with GWAS summary statistics. bioRxiv, 2024.11. 27.625748 (2024).

76. International HapMap, C., et al. Integrating common and rare genetic variation in diverse human populations. Nature 467, 52–8 (2010).

77. Chang, C.C. et al. Second-generation PLINK: rising to the challenge of larger and richer datasets. Gigascience 4, 7 (2015).

78. Bulik-Sullivan, B.K. et al. LD Score regression distinguishes confounding from polygenicity in genome-wide association studies. Nat Genet 47, 291–5 (2015).

79. Su, Z., Marchini, J. & Donnelly, P. HAPGEN2: simulation of multiple disease SNPs. Bioinformatics 27, 2304–5 (2011).

80. Smith, S.M. et al. An expanded set of genome-wide association studies of brain imaging phenotypes in UK Biobank. Nature neuroscience 24, 737–745 (2021).

81. Elliott, L.T. et al. Genome-wide association studies of brain imaging phenotypes in UK Biobank. Nature 562, 210–216 (2018).

82. Beckmann, C.F. & Smith, S.M. Probabilistic independent component analysis for functional magnetic resonance imaging. IEEE transactions on medical imaging 23, 137–152 (2004).

83. Hyvarinen, A. Fast and robust fixed-point algorithms for independent component analysis. IEEE transactions on Neural Networks 10, 626–634 (1999).

84. Zhao, B. et al. Heart-brain connections: Phenotypic and genetic insights from magnetic resonance images. Science 380, abn6598 (2023).

85. Liu, Y. et al. Genetic architecture of 11 organ traits derived from abdominal MRI using deep learning. Elife 10(2021).

86. Sorokin, E.P. et al. Analysis of MRI-derived spleen iron in the UK Biobank identifies genetic variation linked to iron homeostasis and hemolysis. Am J Hum Genet 109, 1092–1104 (2022).

87. Wilman, H.R. et al. Characterisation of liver fat in the UK Biobank cohort. PLoS One 12, e0172921 (2017).

88. Mojtahed, A. et al. Reference range of liver corrected T1 values in a population at low risk for fatty liver disease-a UK Biobank sub-study, with an appendix of interesting cases. Abdom Radiol (NY) 44, 72–84 (2019).

89. Karlsson, A. et al. Automatic and quantitative assessment of regional muscle volume by multi-atlas segmentation using whole-body water-fat MRI. J Magn Reson Imaging 41, 1558–69 (2015).

90. Borga, M. et al. Validation of a fast method for quantification of intra-abdominal and subcutaneous adipose tissue for large-scale human studies. NMR Biomed 28, 1747–53 (2015).

91. West, J. et al. Feasibility of MR-Based Body Composition Analysis in Large Scale Population Studies. PLoS One 11, e0163332 (2016).

92. Linge, J. et al. Body Composition Profiling in the UK Biobank Imaging Study. Obesity (Silver Spring) 26, 1785–1795 (2018).

93. Borga, M. et al. Reproducibility and repeatability of MRI-based body composition analysis. Magn Reson Med 84, 3146–3156 (2020).

94. Langner, T. et al. Kidney segmentation in neck-to-knee body MRI of 40,000 UK Biobank participants. Sci Rep 10, 20963 (2020).

95. Wu, C., Zhang, Z., Yang, X. & Zhao, B. Large-scale imputation models for multi-ancestry proteome-wide association analysis. bioRxiv, 2023.10. 05.561120 (2023).

