## Supplementary material for "PennPRS: a centralized cloud computing platform for efficient polygenic risk score training in precision medicine": supp_figures

**Supplementary Fig. 1: Overview of Online PRS Training Algorithms on PennPRS.**

We developed three single-ancestry methods (C+T-pseudo, Lassosum2-pseudo, and LDpred2-pseudo), two ensemble approaches (Ensemble-pseudo and Ensemble-ARM-pseudo), and one multi-ancestry method (PROSPER-pseudo), all based on pseudo-training without the need for individual-level data. In addition, we have implemented three existing tuning-parameter-free methods (PRS-CS-auto, LDpred2-auto, and DBSLMM) for single-ancestry analysis.

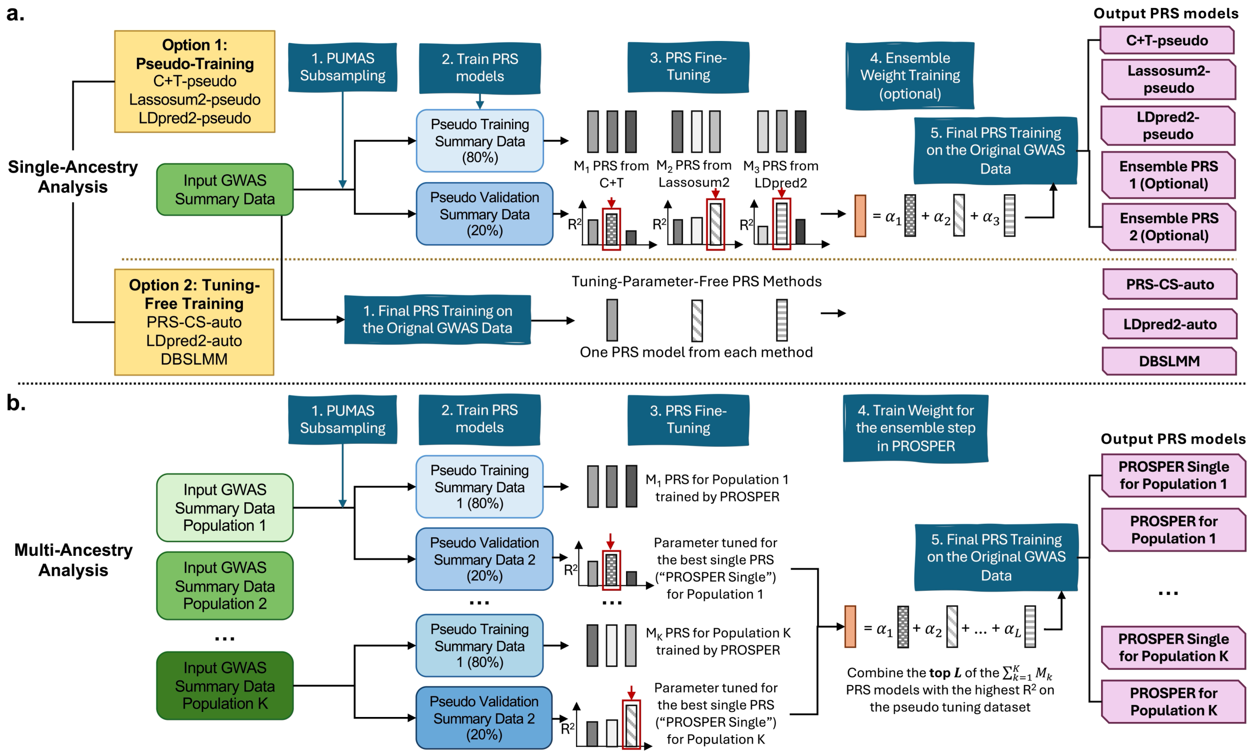

**Supplementary Fig. 2: Comparison of Single-ancestry PRS Pseudo-training and Traditional PRS Methods with Varying Training GWAS Sample Sizes and Sufficient Individual-level Tuning Samples ($N_{tuning}$=2,000).**

We compared the prediction $R^2$ of the PRS trained by C+T-pseudo, Lassosum2-pseudo, LDpred2-pseudo, Ensemble-pseudo, and Ensemble-ARM-pseudo ($R^2_{sum}$) with that of the PRS trained based on individual-level tuning dataset ($R^2_{ind}$) that has a sample size (**a**) $N_{GWAS}$=15K, (**b**) $N_{GWAS}$=45K, or (**c**) $N_{GWAS}$=80K. Results were shown across 10 training GWAS summary datasets and averaged across 100 tuning and testing datasets with $N_{tuning}$ samples for individual data-based parameter tuning and $N_{val}$=2,500 validation samples for calculating prediction $R^2$ for all models. Detailed results are reported in Supplementary Table 1.

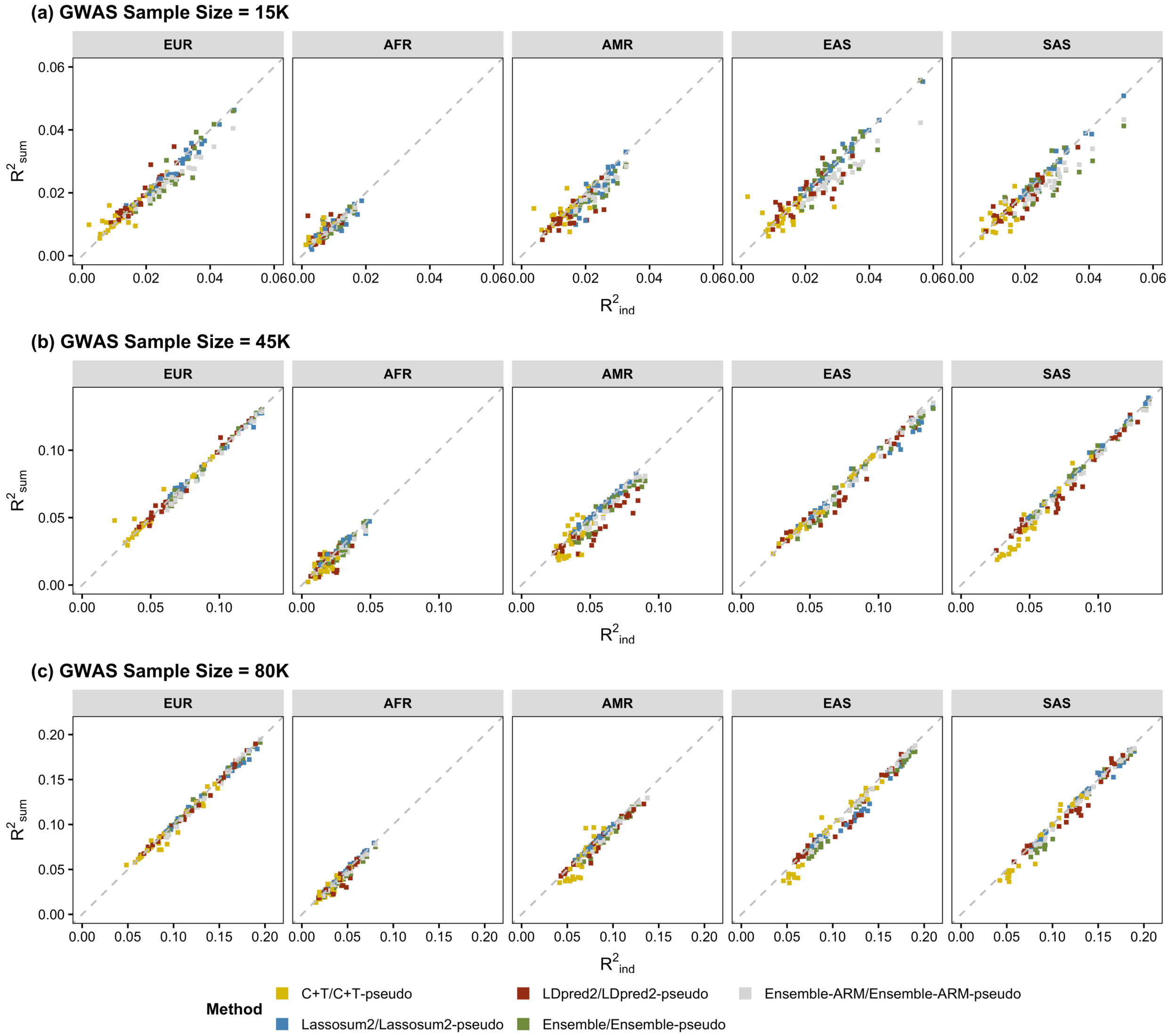

**Supplementary Fig. 3: Comparison of Single-ancestry PRS Pseudo-training and Traditional PRS methods with Individual-level Tuning Data of Various Sample Sizes on brain MRI phenotypes.**

We compared the prediction $R^2$ of the PRS models trained by C+T-pseudo, Lassosum2-pseudo, LDpred2-pseudo, Ensemble-pseudo, and Ensemble-ARM-pseudo ($R^2_{sum}$) with those of PRS models trained based on individual-level tuning dataset ($R^2_{ind}$) that has a sample size $N_{tuning}$ = 1,000, 300, or 100. Results were summarized for 76 resting-state functional MRI (rfMRI), 200 randomly selected diffusion MRI (dMRI), and 200 randomly selected structural MRI (sMRI) traits across 100 random splits, with each split having $N_{tuning}$ tuning samples for individual data-based parameter tuning and the remaining samples as validation samples for calculating prediction $R^2$ for all models.

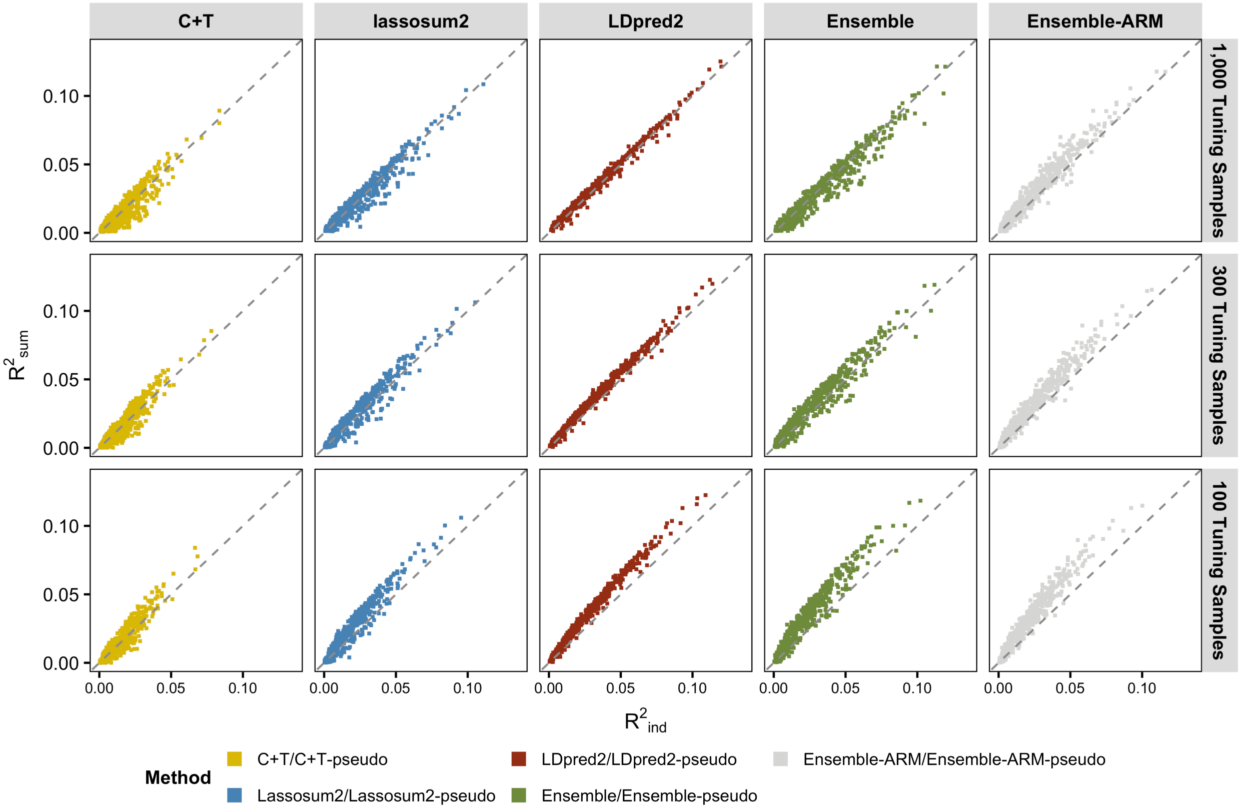

**Supplementary Fig. 4: Evaluation of Multi-Ancestry PRS Pseudo-training on Brain MRI Phenotypes.**

We compared the prediction $R^2$ of the PRS models trained by our proposed pseudo-training approach ($R^2_{sum}$, PROSPER-Single-pseudo and PROSPER-pseudo) with that of the PRS trained based on individual-level tuning datasets ($R^2_{ind}$, PROSPER-Single and PROSPER). Results are summarized by ancestral populations. **a**. Results on 382 brain diffusion MRI (dMRI) phenotypes. The PRS models were trained based on GWAS summary statistics of EUR ancestry from the UK Biobank (UKB) study ($N_{GWAS}$=28,626-32,744) and of EAS ancestry from the CHIMGEN study ($N_{GWAS}$=7,058). Performance was evaluated on hold-out independent UKB samples of EUR ($N_{val}$=4,955) and EAS ($N_{val}$=413-444) ancestries. **b**. Results on 1,181 brain structural MRI (sMRI) phenotypes. The PRS models were trained based on GWAS summary statistics of EUR ancestry from the UKB study ($N_{GWAS}$=30,462-32,646) and of EAS ancestry from the CHIMGEN study ($N_{GWAS}$=7,058). Performance was evaluated on hold-out independent UKB samples of EUR ($N_{val}$=5,050) and EAS ($N_{val}$=426-444) ancestries. For all analyses, we used half of the validation samples for individual-level parameter tuning and the remaining half to report prediction $R^2$ for both our pseudo-training approach and the individual-level tuning data-based training approach. Detailed data information and results are reported in Supplementary Tables 13-14.

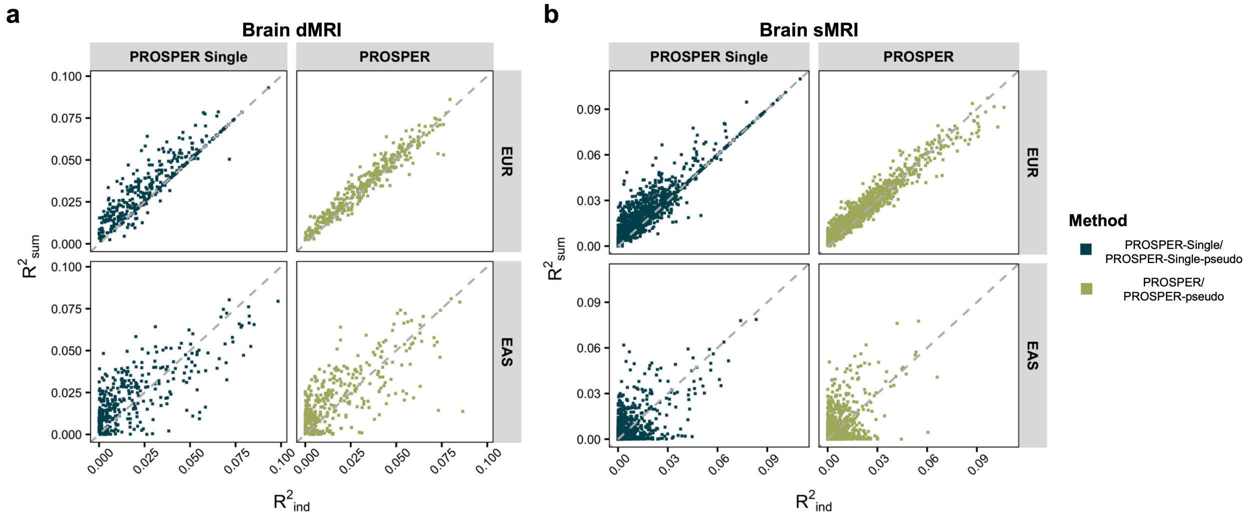

**Supplementary Fig. 5: Comparison between PROSPER-Single-pseudo PRS and PROSPER-pseudo PRS on Blood Lipids and Brain MRI Phenotypes.**

We compared the prediction $R^2$ of the PRS models trained by PROSPER-Single-pseudo with that of PROSPER-pseudo. **a**. Results on 4 blood lipids separately in EUR, AFR, AMR, EAS, and SAS populations. The PRS models were trained based on ancestry-stratified GWAS summary statistics of EUR, AFR, AMR, EAS, and SAS ancestries from the GLGC study ($N_{GWAS}$ =33,658-930,671) and evaluated on the UK Biobank (UKB) samples ($N_{val}$ =750-19,030). **b**. Results on 382 brain diffusion MRI (dMRI) phenotypes separately in EUR and EAS populations reported on UKB samples of EUR ($N_{val}$ =4,955) and EAS ($N_{val}$ =413-444) ancestries, with GWAS summary statistics of EUR ancestry from the UKB study ($N_{GWAS}$=28,626-32,744) and EAS ancestry from the CHIMGEN study ($N_{GWAS}$=7,058). **c**. 1181 brain structural MRI (sMRI) phenotypes separately in EUR and EAS populations reported on UKB samples of EUR ($N_{val}$ =5,050) and EAS ($N_{val}$ =426-444) ancestries, with GWAS summary statistics of EUR ancestry from the UKB study ($N_{GWAS}$=30,462-32,646) and EAS ancestry from the CHIMGEN study ($N_{GWAS}$=7,058). For all analyses, we used half of the validation samples for individual-level parameter tuning and the remaining half to report prediction $R^2$ for both our pseudo training approach and the individual-level tuning data-based training approach. Detailed data information and results are reported in Supplementary Tables 11-14.

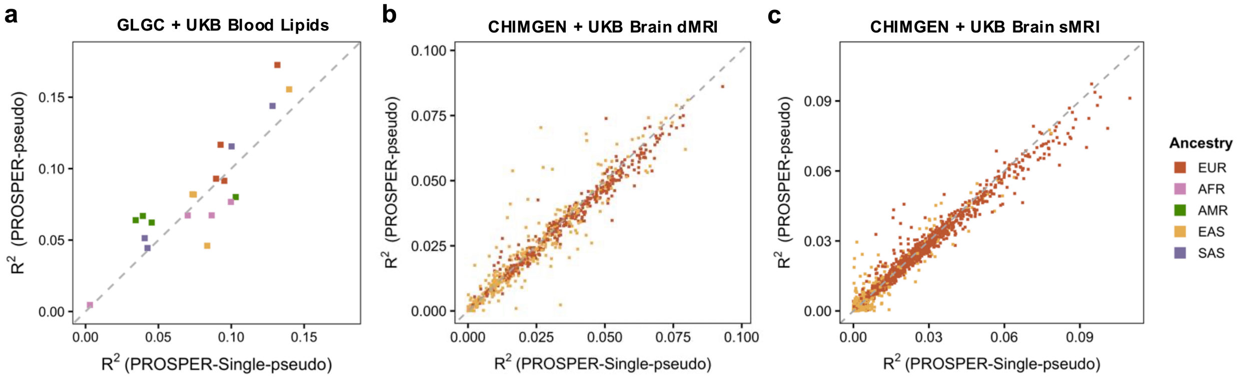
